# Deciphering causal relationships between cell type-specific genetic factors and brain imaging-derived phenotypes and disorders

**DOI:** 10.1101/2024.08.30.24312836

**Authors:** Anyi Yang, Xingzhong Zhao, Yucheng T. Yang, Xing-Ming Zhao

## Abstract

The integration of expression quantitative trait loci (eQTLs) and genome-wide association study (GWAS) findings to identify causal genes aids in elucidating the biological mechanisms and the discovery of potential drug targets underlying complex traits. This can be achieved by Mendelian randomization (MR), but to date, most MR studies investigating the contribution of genes to brain phenotypes have been conducted on heterogeneous brain tissues and not on specific cell types, thus limiting our knowledge at the cellular level. In this study, we employ a MR framework to infer cell type-specific causal relationships between gene expression and brain-associated complex traits, using eQTL data from eight cell types and large-scale GWASs of 123 imaging-derived phenotypes (IDPs) and 26 brain disorders and behaviors (DBs). Our analysis constructs a cell type-specific causal gene atlas for IDPs and DBs, which include 254 and 217 potential causal cell type-specific eQTL target genes (eGenes) for IDPs and DBs, respectively. The identified results exhibit high cell type specificity, with over 90% of gene-IDP and 80% of gene-DB associations being unique to a single cell type. We highlight shared cell type-specific patterns between IDPs and DBs, characterize the putative causal pathways among cell type-specific causal eGenes, DBs and IDPs, and reveal the spatiotemporal expression patterns of these cell type-specific causal eGenes. We also demonstrate that cell type-specific causal eGenes can characterize the associations between IDPs and DBs. In summary, our study provides novel insights into the genetic foundations at the cellular level that influence brain structures, disorders and behaviors, which reveals important implications for therapeutic targets and brain health management.

## Introduction

Genome-wide association studies (GWASs) have identified thousands of variants associated with human brain-associated complex traits, the majority of which are located in non-coding regions[1]. To better understand the biological functions of non-coding genetic signals, expression quantitative trait loci (eQTL), which provide direct links between variants and gene expression levels, have emerged as powerful tools for the interpretation of GWAS loci. High overlap between GWAS loci and eQTLs indicates that certain genetic variants are associated with a higher risk of brain illnesses through the regulation of gene expression[2; 3]. Accordingly, the integration of eQTLs and GWAS findings for identifying causal genes aids in elucidating the biological mechanisms and nominating potential drug targets underlying complex traits. Typically, this can be achieved by Mendelian randomization (MR), an analytic method coping with the challenge of identifying causality, and it has been a vital workflow for understanding the causality between gene expression and complex neurological, psychological, and behavioral traits[4–7].

Previously, studies investigating causal genes for complex traits were largely done by integrating GWAS results with bulk tissue-level eQTLs[5; 8; 9]. However, it has been shown that cell type-level eQTLs affect more constrained genes and have larger effect sizes than tissue-level eQTLs[10]. Several eQTL-related analyses, such as OneK1K[11], Julien et al[10], and MetaBrain[12], have been published to aid in dissecting eQTLs into specific cell types, and have revealed substantial cell type-specific effects on the genetic control of gene expression. Furthermore, studies on single-cell eQTL mapping have shown novel cell type-specific genetic controls of brain disorders which are masked in bulk tissue-level analyses[10; 11; 13; 14]. For example, most genetic variants associated with the risk of Alzheimer’s disease (AD) could potentially act through *cis*-eQTLs in microglia, in contrast with the more polygenic architecture observed in schizophrenia (SCZ), where signals were detected across a broader range of cell types[10]. These studies uncovered that regulation at the cellular level is more responsive to the pathogenic mechanisms of complex disorders than regulation at the bulk level, but our knowledge of how genetic variants contribute to the causal genes of brain traits at the cell type level is limited. Consequently, this has motivated us to identify causal genes in a cell type-specific manner, which will propel us a step forward toward molecular and cellular underpinnings across a wide spectrum of complex traits.

Previous studies have implicated the shared genetic and cellular evidence between brain imaging-derived phenotypes (IDPs) and disorders and behaviors (DBs). High genetic correlation between complex traits and disorders and imaging features has been reported[15; 16]. Recent large-scale GWASs have identified a diverse set of genetic loci and genes linked to both brain imaging phenotypes and various disorders[16–19]. Brain cell type-specific expression has been shown for the causal genes shared among psychiatric, neurodegenerative, and brain structural traits. For example, the causal gene *AKT3*, which is shared among cortical surface area, Parkinson’s disease and major depressive disorder, showed cell type-specific expression in astrocytes[15]. The causal gene *DIP2B*, which is involved in both SCZ and brain stem volume, exhibited cell type-specific expression in excitatory neurons[15]. Common cell type-specific heritability enrichment was identified for the genome-wide SNP variance in both IDPs and DBs. For example, heritability enrichment in oligodendrocytes has been identified in both white matter microstructures and depression[18; 20]. Heritability enrichment in excitatory neurons has been identified in both total surface area[21] and schizophrenia[22]. In light of these findings, we hypothesized that shared genetic underpinnings operating at the cellular level could influence the relationships between brain structural phenotypes and brain disorders, and thus characterizing the underlying putative biological pathways could fuel the development of valuable therapeutic targets.

In this study, we sought to systematically identify putative causal genes that could influence brain structural phenotypes, disorders and behaviors in a cell type-specific manner. We utilized recent *cis*-eQTL data from eight distinct brain cell types, alongside findings from large-scale GWAS of 26 DBs (8 behavioral-cognitive phenotypes, 10 psychiatric disorders and 8 neurological disorders), and 123 IDPs (101 brain regional volumes and 22 white matter tracts). We then applied a MR framework with several complementary approaches, and systematically inferred causal relationships between gene expression levels and these complex phenotypes based on genetic variants. For the genes implicated in these brain-associated complex traits, we further determined the shared cell type-specific eQTL target genes among IDPs and DBs, explored their gene expression patterns using external single-cell data, and characterized the potential causal biological pathways amongst them. Our findings elucidated how genetic variants influence gene expression and further regulate brain structural phenotypes, disorders, and behaviors in specific cell types, which can offer mechanistic insights into healthy and pathological status of the human brain.

## Results

### Overview of the study

Dysregulation of gene expression in specific brain cell types has been widely reported in psychiatric and neurological disorders[10; 11; 13; 14; 23]. To explore the potential causal relationships between genetic variants and diverse brain-associated complex traits in a cell type-specific manner, we performed two-sample MR analysis by integrating GWAS summary statistics of 26 DBs (**Supplementary Table 1**) and 123 IDPs (**Supplementary Table 2**) with a recently published cell type-specific *cis*-eQTL catalog from eight brain cell types[10] (**Supplementary Table 3**). The brain cell types included astrocytes, endothelial cells, excitatory neurons, inhibitory neurons, microglia, oligodendrocytes, oligodendrocyte precursor cells, and pericytes. Given the existing evidence of bidirectional causal influence between brain disorders and brain imaging-derived phenotypes[24; 25], we selected 26 DBs, which can be summarized into three groups, i.e., 8 behavioral-cognitive phenotypes, 8 neurological disorders, and 10 psychiatric disorders (GWAS average sample size: 284,168; **Supplementary Table 1**); and also 123 IDPs, which can be summarized into two groups, i.e., 101 brain regional volumes and 22 white matter microstructure phenotypes measured by the mean fractional anisotropy (GWAS average sample size: 22,072; **Supplementary Table 2**). The study design is shown in **Fig. 1a**. We examined the potential associations between gene expression levels and these brain-associated complex traits via a transcriptome-wide association study (TWAS)[26], and retained a set of 43,494 gene-DB pairs and 284,927 gene-IDP pairs with nominal *P* values < 0.05 for subsequent MR analyses. To ensure that the instrumental variables (IVs) were valid, only the single nucleotide polymorphisms (SNPs) with genome-wide significance (*P* < 5×10^-8^) were selected as IVs. We also manually checked the sample description of the *cis*-eQTL catalog and each GWAS, and confirmed the lack of overlap between these datasets.

**Fig. 1.**
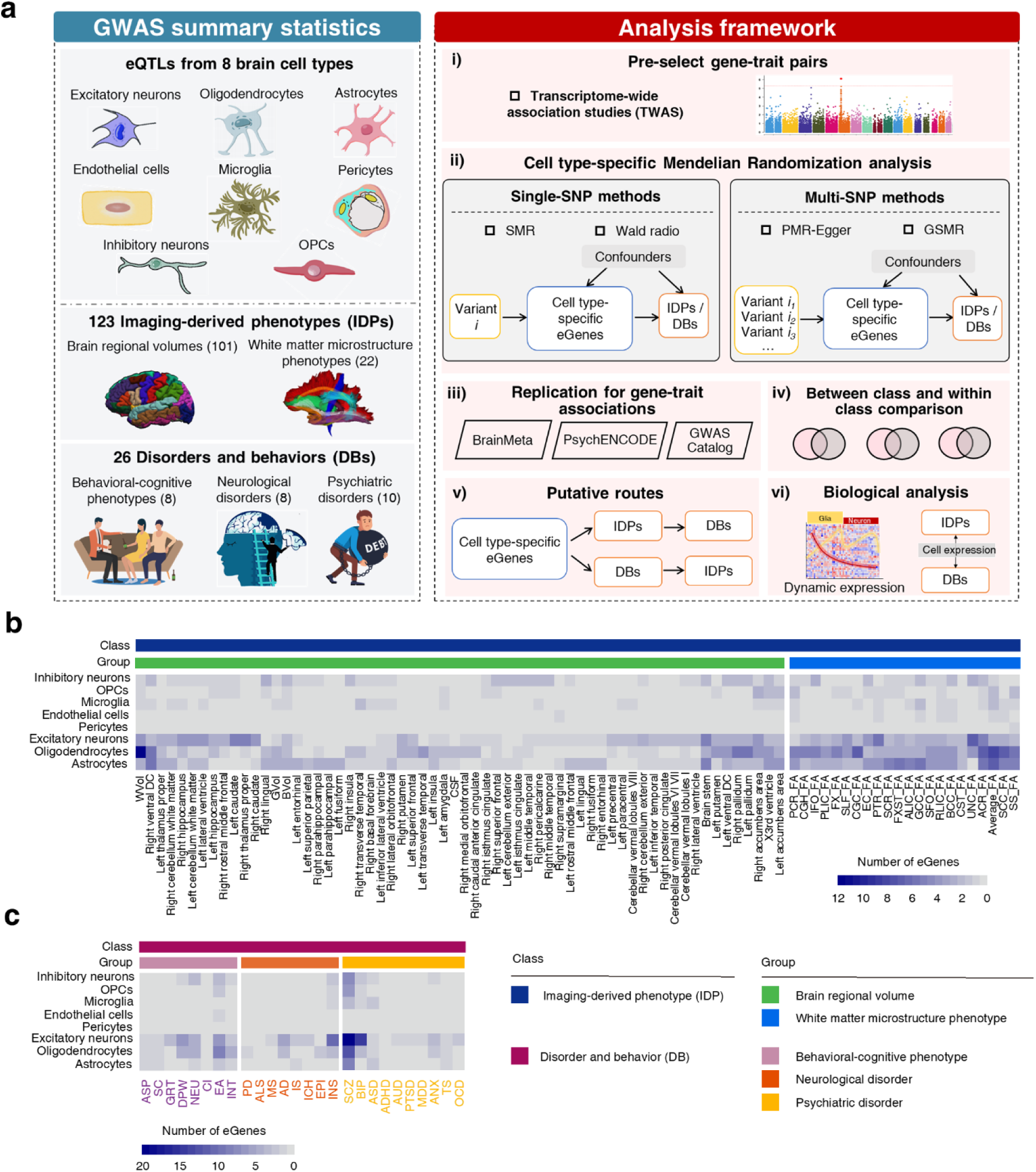
Study workflow and count of significant cell type-specific causal eGenes across brain-associated complex traits. (a) Study workflow of our analysis. We leveraged eQTL data from eight cell types and large-scale GWASs of 123 IDPs and 26 DBs. After pre-selecting potential associated gene-trait pairs using a transcriptome-wide association study, we employed a cell type-specific MR framework to infer putatively causal eGenes of these 149 brain-associated complex traits (categorized into two classes of DBs and IDPs, and five groups of psychiatric disorders, neurological disorders, behavioral-cognitive phenotypes, brain regional volumes, and white matter tracts). Further downstream analysis included determining shared cell type-specific eQTL target genes among IDPs and DBs, exploring their expression patterns using single-cell data, and characterizing the potential causal pathways amongst them. The clip-art of brain regional volumes is from the Desikan-Killiany Atlas[165], and the clip-art of white matter tracts is from the Atlas Track[166]. **(b)** Heatmap showing the number of putatively causal cell type-specific eGenes across IDPs. Only the IDPs that had more than three putatively causal cell type-specific eGenes are shown in this figure. **(c)** Heatmap showing the number of putatively causal cell type-specific eGenes across DBs.

To ensure the robustness of the MR inference for each cell type, we employed four different MR methods, including SMR[4], Wald radio[27], PMR-Egger[6] and GSMR[28]. These methods rely on complementary analytical frameworks, with different strategies for selecting IVs (i.e., one-SNP or multi-SNP; independent SNPs or correlated SNPs), MR assumptions (i.e., permitting horizontal pleiotropy or not), and targeting exposures (i.e., a universal method or a method specifically tailored for gene expression as the exposure) (**Methods**; **Supplementary Fig. 1**). We tested the reliability of the predicted causal eGene-trait pairs (i.e., the predicted cell type-specific eQTL target genes implicated in diverse brain-associated complex traits) using additional functional genomic datasets from the human brain. We analyzed the trait pleiotropy of these putatively causal cell type-specific eGenes (denoted as cell type-specific causal eGenes hereinafter) across different classes and groups of these brain-associated complex traits. Finally, we reconstructed putative causality routes among cell type-specific causal eGenes, DBs and IDPs, and characterized the spatiotemporal expression patterns of the cell type-specific causal eGenes implicated in DBs and IDPs.

### Putative causal effects of cell type-specific eGenes on IDPs and DBs

As shown in **Fig. 1b** and **Supplementary Table 4**, we identified 254 eGenes that may have causal effects on 112 IDPs in eight cell types (FDR < 0.05), forming a catalog of 760 combinations between cell types, eGenes and IDPs. Among these, 184 and 112 eGenes were inferred to have causal effects on 90 brain regional volumes and 22 white matter microstructure phenotypes, respectively. To confirm the reproducibility of the predicted eGene-IDP pairs, we performed MR analyses using cortical *cis*-eQTL summary statistics from BrainMeta[29], and found that 76.3% (505 out of 662; *P* < 10^-16^, hypergeometric test) of the eGene-IDP pairs could be successfully replicated (**Supplementary Table 4** and **Supplementary Fig. 2**). Our predicted eGene-IDP pairs exhibited strong cell type specificity, with 91.4% (605 out of 662) existing only in a particular cell type, which is consistent with previous literature showing that the formation of brain structures is modulated by genes in a cell type-specific manner[30–32]. White matter microstructure phenotypes tended to exhibit an overall greater number of causal eGenes in oligodendrocytes and astrocytes than brain regional volumes did (**Fig. 1b**), which aligns with previous studies showing that white matter microstructure shows significant heritability enrichment in glial cells such as oligodendrocytes and astrocytes[18]. The pair between white matter volume and oligodendrocytes exhibited the most causal eGenes (12 eGenes), eight of which (*ANKRD44*, *ZCWPW1*, *SLC16A8*, *PILRB*, *RUNX2*, *LARP6*, *C7orf61*, and *SUPT3H*) have been reported to be associated with neuroimaging measurements in the GeneCards database[33].

As shown in **Fig. 1c** and **Supplementary Table 5**, we identified 217 eGenes that may have causal effects on 26 DBs in eight cell types (FDR < 0.05), forming a catalog of 298 combinations between cell types, eGenes and DBs. Among these, 109, 61 and 85 eGenes were inferred to have a causal effect on 10 psychiatric disorders, 8 neurological disorders and 8 behavioral-cognitive phenotypes, respectively. As we did for the IDPs, we confirmed the reproducibility of eGene-DB pairs using three distinct datasets: the cortical *cis*-eQTL summary statistics from BrainMeta[29], differentially expressed genes and transcripts from PsychENCODE[23], and documented associations between genes and complex traits from GWAS Catalog[34] (see **Methods**). We found that 52.9% (127 out of 240; *P* < 10^-16^, hypergeometric test), 29.5% (23 out of 78; *P* < 2×10^-4^, hypergeometric test) and 23.8% (57 out of 240; *P* < 10^-16^, hypergeometric test) of the eGene-DB pairs were successfully replicated, respectively (**Supplementary Table 5** and **Supplementary Fig. 2**). Consistent with our observations for IDPs, the predicted eGene-DB pairs were highly cell type specific, with 82.1% (197 out of 240) existing only in a particular cell type. These observations suggest that most of the genes implicated in brain-associated disorders and behaviors generally function through a particular cell type, which is in agreement with previous studies[10; 35]. Excitatory neurons displayed the most pronounced number of causal eGenes across almost all trait groups, particularly in SCZ, BIP from the psychiatric disorder group, and INS from the neurological disorder group. In contrast, inhibitory neurons exhibited fewer causal eGenes than excitatory neurons but still demonstrated notable involvement in disorders like SCZ. The pair between SCZ and excitatory neurons exhibited the most causal eGenes (20 eGenes), four of which (*ANKRD27*, *LIN28B*, *UROS*, and *CCHCR1*) have been reported to be associated with SCZ in the GeneCards database[33]. Almost all the eGenes that were predicted to be causally associated with AD in excitatory neurons (5 out of 6 eGenes) have been reported to be implicated in AD and other neurodegenerative disorders, such as *MSH3*[36], *ICA1L*[37], *RGS14*[38], *C17orf97*[39] and *ZSCAN31*[40], suggesting their common and cumulative roles in the pathogenesis of neurodegenerative disorders.

### Pervasive trait pleiotropy of causal eGenes

To explore the trait pleiotropy of these putative causal eGenes, we then systematically compared the causal eGenes in terms of their cell type-specificity across different trait classes (i.e., IDP and DB) and their corresponding trait groups (i.e., brain regional volumes and white matter microstructure phenotypes belonging to the IDP class; behavioral-cognitive phenotypes, neurological disorders and psychiatric disorders belonging to the DB class). We observed significant overlap of the causal eGenes between each pair of trait groups (all with *P* < 10^-10^, hypergeometric test; **Supplementary Fig. 3**). In terms of within-class trait pleiotropy, the brain regional volume group and white matter microstructure phenotype group showed the greatest overlap (a total of 42 shared causal eGenes [37.5%]), in which the strongest overlap with the brain regional volume group among the individual white matter microstructure phenotypes was from the inferior fronto-occipital fasciculus (IFO) ([22.81%]; **Fig. 2a** and **Supplementary Fig. 4**). For the DB class, four causal eGenes — *DDHD2* in excitatory neurons, *MAP2K5* in inhibitory neurons, *XKR6* in inhibitory neurons, and *LIN28B* in excitatory neurons — were found to be mutually causal across the neurological disorder, behavioral-cognitive phenotype and psychiatric disorder groups (**Supplementary Table 6**). These genes have been reported as potential causatives for multiple psychiatric brain disorders[41–44].

**Fig. 2.**
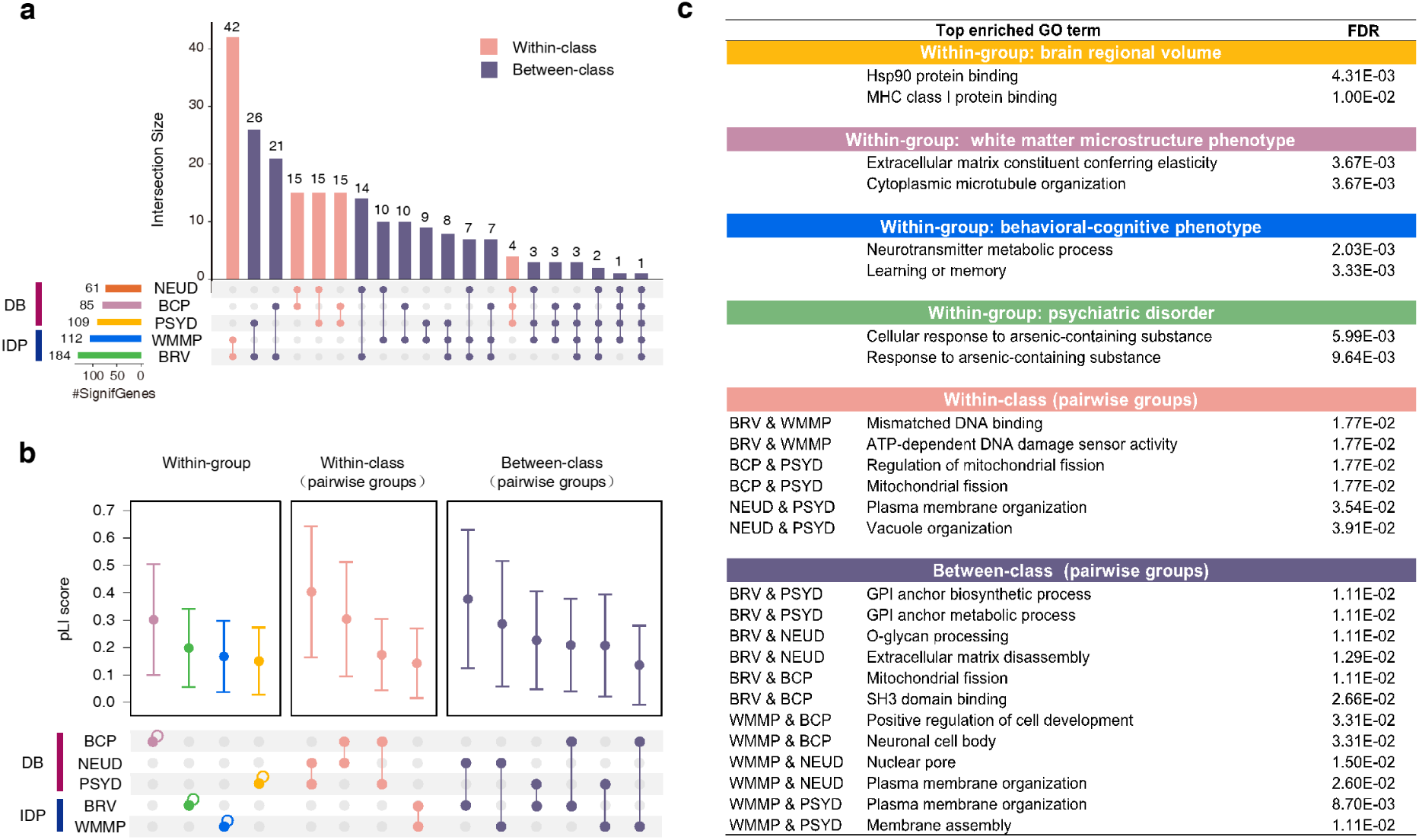
Putative causal genes shared among IDPs and DBs. (a) UpSet figure showing the identified cell type-specific causal eGenes that are mutually shared across trait classes and groups. **(b)** The probability of loss-of-function intolerance (pLI) scores for causal eGenes shared across trait classes and groups (pairwise sharing). **(c)** The most enriched gene sets (top two) of the genes shared across trait groups and classes (pairwise sharing). Within-group: genes shared by at least two traits within each group. Within-class: genes shared by at least one trait in a group of a class and at least one trait in another group of the same class. Between-class: genes shared by at least one trait in the DB class and another trait in the IDP class. BRV: Brain regional volume; WMMP: White matter microstructure phenotype; BCP: Behavioral-cognitive phenotype; NEUD: Neurological disorder; PSYD: Psychiatric disorder.

In terms of between-class trait pleiotropy, the greatest overlap was shown between the brain regional volume group and behavioral-cognitive phenotype group (a total of 21 shared causal eGenes [24.7%]), in which the strongest overlap with the brain regional volume group among the individual behavioral-cognitive phenotypes was from drinks per week (DPW) ([10.45%]; **Fig. 2a** and **Supplementary Fig. 4**); and between the brain regional volume group and psychiatric disorder group (a total of 26 shared eGenes [23.85%]), in which the strongest overlap with the brain regional volume group among the individual psychiatric disorders was from ADHD ([15.38%]; **Fig. 2a** and **Supplementary Fig. 4**). We also detected many causal eGenes shared among three or more groups from different trait classes (**Fig. 2a** and **Supplementary Table 6**). Specifically, 17 causal eGenes were found as shared among two IDP groups and one DB group, nine causal eGenes were shared among one IDP group and two DB groups, four causal eGenes were shared among four groups (*XKR6* in inhibitory neurons, *MAPT* in astrocytes, *ZSCAN31* in astrocytes, *MSH3* in excitatory neurons), and one causal eGene was found as shared among all the five trait groups (*XKR6* in inhibitory neurons). Overall, these findings suggest a shared genetic susceptibility specific to cell types across different groups of brain-associated complex traits.

The probability of loss-of-function intolerance (pLI) score is a genetic constraint metric that quantifies the tolerance of a given gene to loss-of-function mutations. To further depict the biological implications of the causal eGenes that were shared between brain-associated complex traits, we then characterized the gene conservation using pLI scores from ExAc[45], as well as performed gene-set enrichment analysis[46]. In terms of the shared causal eGenes between pairs of traits within each group (left panel in **Fig. 2b**), the behavioral-cognitive phenotype group showed the highest pLI scores (*μ* = 0.302, *σ^2^* = 0.203), followed by the brain regional volume group (*μ* = 0.20, *σ^2^* = 0.14). Further functional enrichment analysis revealed that the causal eGenes shared within the behavioral-cognitive phenotype group were enriched in biological pathways associated with memory, cognition, and neurotransmitters (**Fig. 2c** and **Supplementary Table 7**), which is consistent with previous studies highlighting their critical roles in regulating learning, memory formation, and cognitive performance[47; 48]. In contrast, the causal eGenes shared within the brain regional volume group were generally involved in protein binding, such as Hsp90 protein binding and MHC class I protein binding (**Fig. 2c** and **Supplementary Table 7**), which aligns with previous findings showing that the expression of Hsp90 and MHC class I proteins could regulate synapse formation and brain development[49; 50].

Next, we explored the conservation and enriched biological functions of the shared causal eGenes between pairs of traits within the same trait class (middle panel in **Fig. 2b**). A greater level of genetic conservation was observed among shared causal eGenes within the DB class than within the IDP class. For example, the causal eGenes shared between the neurological disorder and psychiatric disorder groups demonstrated the highest pLI scores (*μ* = 0.409, *σ^2^* = 0.235), in contrast to those shared by the brain regional volume and WMM groups, which presented the lowest scores (*μ* = 0.153, *σ^2^* = 0.125). This observation indicates that the genes implicated in diverse brain disorders are more conserved than those implicated in brain structural phenotypes[51]. Consistently, the gene-set enrichment analysis revealed that the causal eGenes shared between the neurological disorder and psychiatric disorder groups were involved in biological functions, such as cell maintenance and cell signaling (e.g., plasma membrane organization; **Fig. 2c** and **Supplementary Table 7**), which have been linked to the etiology of neuropsychiatric disorders[52; 53].

Similarly, we further analyzed the conservation and enriched biological functions for the shared causal eGenes between pairs of traits from different classes (right panel in **Fig. 2b**). Notably, the genes shared between the brain regional volume group and groups in the DB class (i.e., the behavioral-cognitive phenotype, neurological disorder and psychiatric disorder groups) showed higher pLI scores than those shared between the white matter microstructure phenotype group and the groups from the DB class, suggesting that, compared with white matter microstructure, volumetric measures may capture more fundamental and evolutionarily conserved genetic factors underlying these brain disorders and behaviors. These observations align with findings from prior studies that brain microstructural changes typically precede volumetric changes in the progression of brain disorders and behaviors[54; 55], further suggesting that the genetic factors influencing these early microstructural changes might be less evolutionarily conserved. Additionally, the causal eGenes shared between the brain regional volume and neurological disorder groups, which displayed the highest pLI scores between trait classes, were significantly enriched with four biological pathways related to extracellular matrix dynamics (**Fig. 2c** and **Supplementary Table 7**). The extracellular matrix is crucial for shaping brain volume, nervous system development and maintenance, and the pathophysiology of brain disorders[56–58], and our findings highlight the importance of extracellular matrix regulation in the shared genetic basis of brain volume and neurological disorders.

### Shared cell type specificity between different trait classes

After elucidating the genetic characteristics shared across brain-associated complex traits, we focused on the common cell types in which the causal eGenes were from two trait classes: IDPs and DBs. We observed substantial differences in the brain cell types for the causal eGenes that were shared between the DB and IDP classes (**Fig. 3a**). It was revealed that a broad range of cell types were shared between SCZ and the brain regional volume group, including astrocytes, excitatory neurons, microglia, oligodendrocytes, and OPCs; as well as between DPW and the brain regional volume group, including astrocytes, excitatory neurons, microglia, inhibitory neurons, and OPCs (**Fig. 3a**). This finding is consistent with the polygenic architecture of SCZ and DPW[59; 60]. In contrast, neurodevelopmental disorders such as ASD, PTSD, and ADHD exhibited restricted cell type (i.e., inhibitory neurons) that were shared with the brain regional volume and white matter microstructure phenotype groups (**Fig. 3a**). This finding supports the hypothesis that abnormalities in neurodevelopmental disorders may stem from disrupted neural communication within inhibitory neurons, thereby affecting brain structure and function[61–63]. Risky behaviors such as general risk tolerance (GRT), automobile speeding propensity (ASP), and DPW also exhibited significant sharing with the brain regional volume and white matter microstructure phenotype groups within inhibitory neurons (**Fig. 3a**). Since the literature has suggested the influence of region-specific inhibitory activity of neurons on risky behaviors[64], we further looked at the specific brain regions that significantly share cell type-specific causal eGenes with these risky behaviors. In particular, we found that these regions were generally involved in executive and visual functions, including fornix-stria terminalis (FXST), IFO, uncinate fasciculus (UNC), corona radiata, orbitofrontal cortex, isthmus cingulate, and ventral diencephalon (**Supplementary Fig. 5**)[65–73]. Our results suggest a genetic link between risky behaviors and brain regions related to executive and visual functions in inhibitory neurons. Furthermore, only the traits in the behavioral-cognitive phenotype group (i.e., DPW, ASP and educational attainment) and SCZ exhibited a significant overlap of shared causal eGenes with the brain regional volume group in OPCs (**Fig. 3a**). This supports previous literature suggesting that gene expression changes in OPCs are associated with critical stages of brain development and affect the formation of neural circuits involved in behavioral and cognitive outcomes[74; 75].

**Fig. 3.**
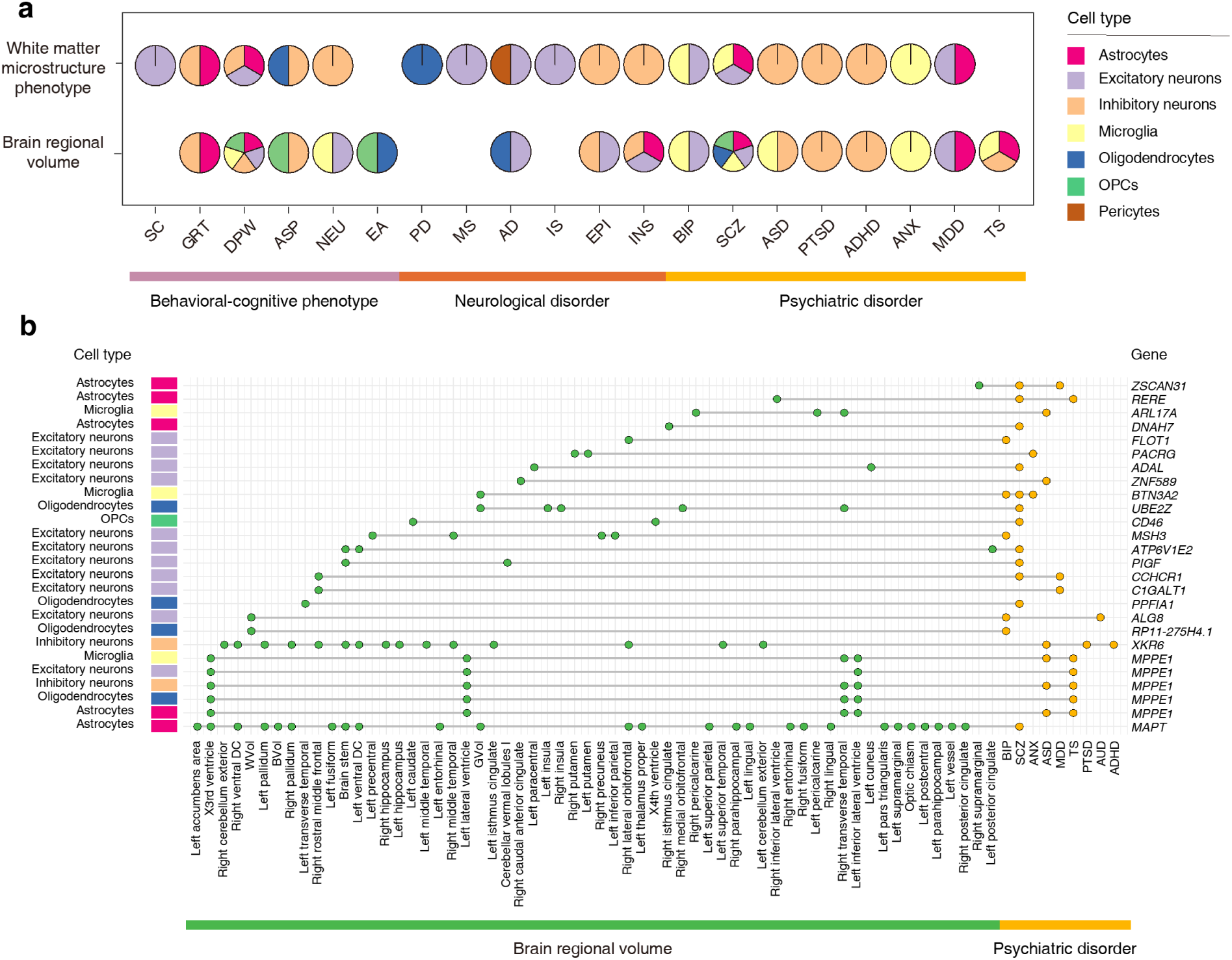
Potentially shared cell type-specific patterns between IDPs and DBs. (a) Pie plot showing the brain cell types where cell type-specific causal eGenes were significantly shared between the IDP groups and each individual trait from the DB class. The hypergeometric test was used to test the sharing of cell type-specific causal eGenes within a specific cell type. Significant brain cell types were identified at the significance threshold of false discovery rate (FDR) < 0.05. **(b)** Cell type-specific causal eGenes shared between brain regional volumes and psychiatric disorders.

Our findings pointed to a common cell type-specific genetic susceptibility between brain structures and these brain disorders and behaviors. We first explored the cell type-specific causal eGenes shared between IDPs and psychiatric disorders. The *MAPT* gene in astrocytes showed pronounced sharing across SCZ, 26 brain regional volumes, and 15 white matter microstructure phenotypes (**Fig. 3b** and **Supplementary Fig. 6**). Our analysis associated increased expression of *MAPT* in astrocytes with an elevated risk of SCZ, as well as with reduced mean FA in 14 white matter tracts and decreased volume in 17 brain regions. The *MAPT* gene is located at 17q21 and is known for its pivotal role in human brain size and patterning, which is related to multiple brain imaging features like intracranial volume, white matter microstructure and cerebral cortex structure[17]. Studies have shown that *MAPT* mutations can influence brain atrophy[76], and that dysregulation of *MAPT* gene expression is linked to SCZ[77]. Additionally, a strong causal contribution of glial pathology, including astrocytic dysfunction, to SCZ was highlighted previously[78]. Crucially, this gene encodes the tau protein, which is essential for stabilizing microtubule architecture and facilitating axonal transport in neurons[79]. Given the central role of tau in both neuronal and glial cells, coupled with existing research of the modulation of tau pathology in neurodegenerative disorders[80], our findings support the potential therapeutic value of targeting *MAPT* gene expression in astrocytes for addressing SCZ pathophysiology and related brain abnormalities.

We next explored the cell type-specific causal eGenes shared between IDPs and neurological disorders. The genes *ZSCAN31* in excitatory neurons[40; 81], *SYT14* in excitatory neurons[82; 83], and *ICA1L* in excitatory neurons[37; 84] are shared among AD and multiple IDPs (**Supplementary Figs. 6 and 7**), which is consistent with previous studies showing that the variants located in these genes were implicated in both brain measurements and AD[37; 40; 81-84].

Finally, we explored the cell type-specific causal eGenes shared between IDPs and behavioral-cognitive phenotypes. The *XKR6* gene in inhibitory neurons was shared among the risky behaviors of ASP and GRT, 15 brain regional volumes (including right lateral orbitofrontal, left isthmus cingulate, and left/right ventral diencephalon), and 5 white matter microstructure phenotypes (including the mean FA of FXST, IFO, and UNC) (**Supplementary Figs. 6 and 7**). The *XKR6* gene, also known as XK-related 6, is classified as a member of the Kell blood group complex subunit-related family[85]. A recent large-scale GWAS for risky behavior identified associated variants within the *XKR6* gene, and also suggested that inhibitory neurotransmission may play a role in shaping the variation in general risk tolerance across individuals[86]. Coupled with the observed genetic link between risky behaviors and brain regions related to executive and visual functions in inhibitory neurons (**Fig. 3a** and **Supplementary Fig. 5**), our results imply that the expression level of *XKR6* in inhibitory neurons may affect brain areas related to executive and visual functions; this potentially sensitizes individuals to external visual stimulation and influences their risk-tolerant willingness. Meanwhile, a causal relationship was found between the expression level of the *SRR* gene in astrocytes and smoking cessation (SC), drinks per week (DPW), neuroticism (NEU), 5 brain regional volumes, and one white matter microstructure phenotype. The *SRR* gene, encoding serine racemase, is expressed in excitatory glutamatergic neurons within the human forebrain[87], and plays a key role in glutamatergic synaptic signaling[88]. Prior research has linked the expression of *SRR* to substance use traits and identified *SRR* as a promising drug target for novel smoking treatment strategies[88].

### Putative causality routes among cell type-specific causal eGenes, and brain-associated complex traits

Previous studies have suggested that genetic risk factors may influence complex disorders by impacting the endophenotypes of brain traits[25; 89; 90], or that they may affect complex disorders and behaviors by altering brain structures[25]. Consequently, characterizing the causality routes among cell type-specific causal eGenes, IDPs and DBs could offer valuable insights into the regulatory mechanisms underlying brain structural abnormalities and disorders. We applied an analogical MR pipeline as described above to investigate the bidirectional causal relationships between the abovementioned IDPs and DBs (**Methods**), and identified six causal associations of IDPs on DBs, and three causal associations of DBs on IDPs. By integrating eGene-IDP pairs with eGene-DB pairs, we ultimately constructed four cell type-specific eGene-IDP-DB routes and seven cell type-specific eGene-DB-IDP routes, where the cell type-specific eGene was identified as causal for both IDP and DB in each individual route (**Fig. 4** and **Supplementary Table 8**).

**Fig. 4.**
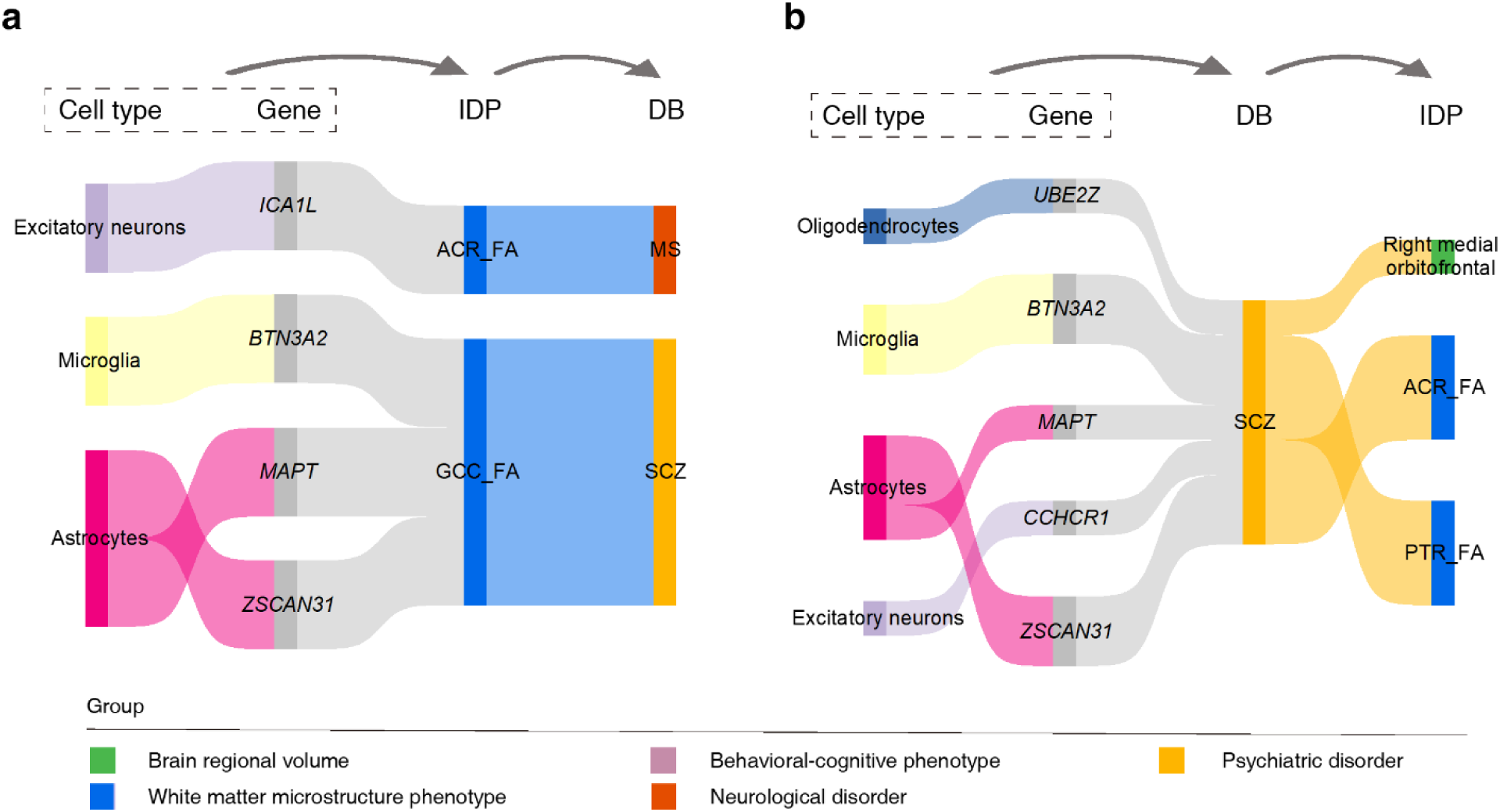
Putative routes among cell type-specific causal eGenes, IDPs, and DBs. (a) Putative cell type-specific causal eGene-IDP-DB routes. In each route, the cell type-specific eGene was identified as causal for both IDP and DB and the IDP was identified as causal for the DB. **(b)** Putative cell type-specific causal eGene-DB-IDP routes. In each route, the cell type-specific eGene was identified as causal for both IDP and DB, and the DB was identified as causal for the IDP. Detailed results can be found in **Supplementary Table 8**.

Here, a putative eGene – IDP – DB causality route was showcased: *ICA1L* in excitatory neurons – anterior corona radiata (ACR) – MS (**Fig. 4a** and **Supplementary Table 8**). The protein-coding gene *ICA1L*, which encodes a member of the interacting BAR-domain family of proteins[91], has been implied in impaired excitatory synaptic signaling[92]. Existing studies have suggested that dysregulation of *ICA1L* at the expression level confers the pathogenesis of MS[93], and that a genetic variant in *ICA1L* is also associated with the microstructure of ACR[16] which functions in executive control[94]. Furthermore, diffusion tensor imaging (DTI) abnormalities of the corona radiata have been reported to be correlated with MS[95]. As indicated by our results, *ICA1L* gene expression in excitatory neurons may affect the white matter microstructure of ACR, leading to abnormalities in executive control, which would increase the risk of MS (ACR to MS: effect size=0.11, FDR=0.002 for PMR-Egger, and effect size=1.06, FDR=21.6×10^-16^ for GSMR); (*ICA1L* on ACR: effect size=-0.16, FDR=5.42×10^-4^ for SMR, effect size=-0.17, FDR=1.16×10^-36^ for Wald Ratio, and effect size=-0.15, FDR=1.91×10^-4^ for GSMR); (*ICA1L* on MS: effect size=-0.2, FDR=0.011 for SMR, effect size = −0.19, FDR = 1.1×10^-6^ for Wald Ratio, and effect size=-0.19, FDR=9.63×10^-3^ for GSMR). Other interesting examples include *MAPT* in astrocytes – genu of corpus callosum (GCC) – SCZ, which indicates that the expression level of *MAPT* in astrocytes may affect the microstructure of GCC and further contribute to SCZ susceptibility[96; 97].

We showcased another putative eGene – DB – IDP route: *ZSCAN31* in astrocytes – SCZ – posterior thalamic radiation (PTR) (**Fig. 4b** and **Supplementary Table 8**). The gene *ZSCAN31* encodes a protein containing multiple C2H2-type zinc finger motifs. The expression of this gene may influence SCZ risk[98], and may be link with neuroimaging measurements[81]. Thalamic radiations are fibers that connect the thalamus and the cerebral cortex. Previously, alterations of thalamic radiation integrity were reported in individuals with SCZ[99]. Moreover, thalamocortical dysconnectivity was suggested to be more prominent in individuals who later develop full-blown psychosis illness, which indicates the progressive nature of abnormal thalamic connectivity in SCZ patients[100]. As indicated by the identified coherent biological route, the expression level of *ZSCAN31* in astrocytes may contribute to the causal effect of SCZ disease progression on the microstructure of PTR (SCZ to PTR: effect size=0.087, FDR=1.54×10^-5^ for PMR-Egger, and effect size=0.033, FDR=1.45×10^-5^ for GSMR); (*ZSCAN31* on SCZ: effect size=0.17, FDR=2.38×10^-26^ for Wald Ratio, effect size =0.14, FDR = 1.53×10^-5^ for GSMR, and effect size=0.16, FDR=6.89×10^-5^ for SMR); (*ZSCAN31* on PTR: effect size=0.08, FDR=2.81×10^-8^ for Wald Ratio, effect size=0.08, FDR=2.57×10^-3^ for GSMR, and effect size=0.08, FDR=0.03 for SMR). In summary, these integrated causality routes hold promise for further elucidating the regulatory mechanisms and potential diagnostic markers of brain illnesses.

### Spatiotemporal expression patterns of causal eGenes for IDPs and DBs

Our MR results motivated us to investigate the expression levels of the causal eGenes for IDPs and DBs in human brain (**Methods**). We found that the causal eGenes of DBs were more highly expressed in glial cells than in neurons (*P* < 2.2×10^-16^, Wilcoxon test), particularly in astrocytes (**Supplementary Fig. 8**). The causal eGenes of psychiatric disorders and behavioral-cognitive phenotypes showed significantly higher expression in neurons during the prenatal period than the postnatal period, whereas the opposite pattern was observed for glial cells (*P* < 0.001, Wilcoxon test; **Fig. 5a** and **Supplementary Fig. 9a**). Most causal eGenes of neurological disorders showed elevated expression in neural cells during the postnatal period. However, the causal eGenes of ALS were exclusively expressed in neural cells, with elevated expression after birth (**Supplementary Fig. 9b**), consistent with previous studies[101–103]. Additionally, we investigated the expression trajectories of the causal eGenes of DBs across the whole lifespan. We found that the expression of causal eGenes of psychiatric disorders decreased until late mid-fetal development (19-24 PCW) and then gradually increased with age until early childhood (1-6 Years) in glial cells, but decreased in neural cells (**Figs. 5b and 5c**, **Supplementary Fig. 10**). As expected, the expression trajectories of the causal eGenes of behavioral-cognitive phenotypes were similar to those of psychiatric disorders (**Supplementary Fig. 11**).

**Fig. 5.**
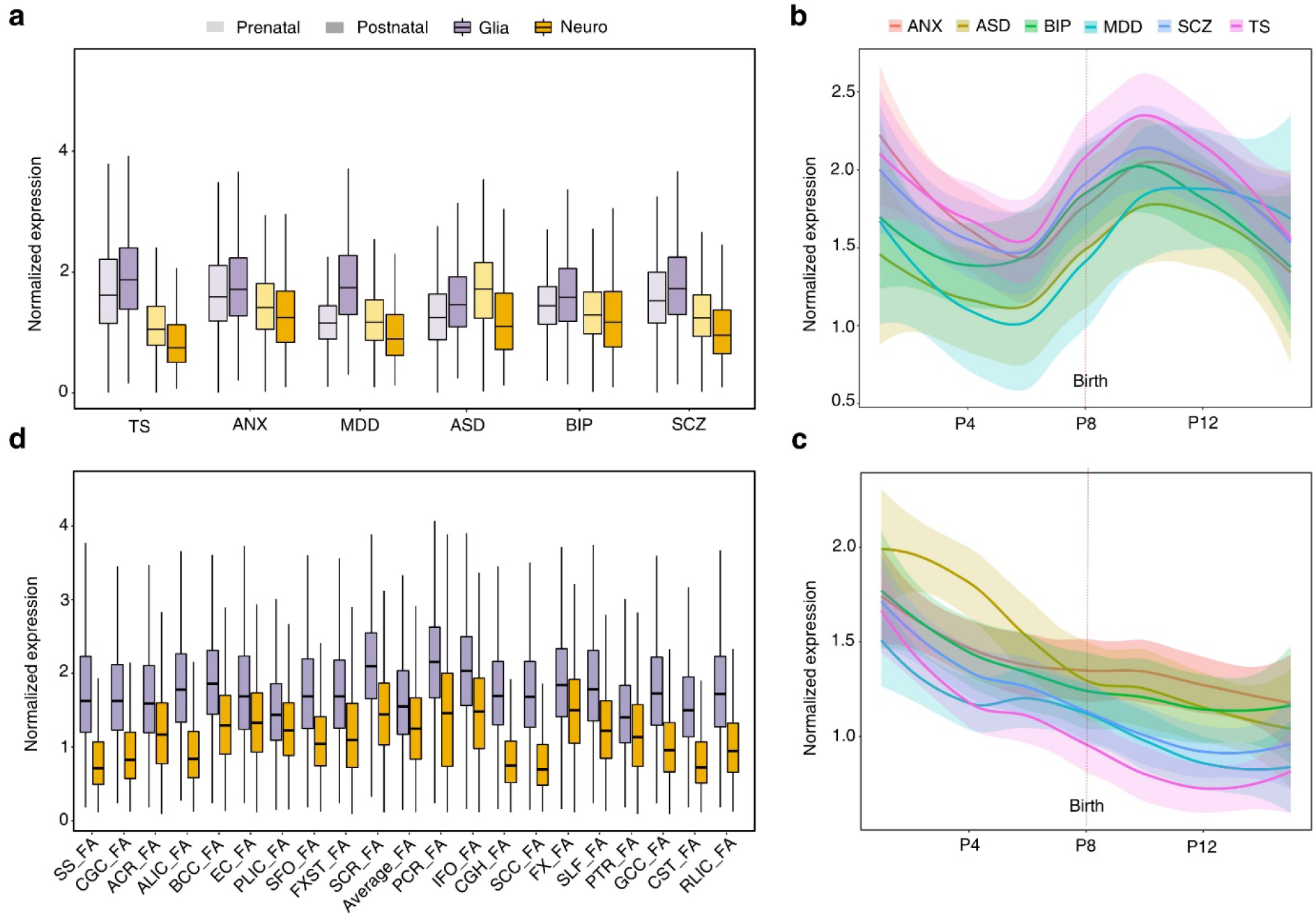
Spatiotemporal expression for causal eGenes of psychiatric disorders and white matter microstructure phenotypes. (a) Expression of psychiatric disorder-associated causal eGenes in the prenatal stage versus the postnatal stage in neural and glial cell types. **(b)** Expression dynamics of psychiatric disorder-associated causal eGenes in glial cells. **(c)** Expression dynamics of psychiatric disorder-associated causal eGenes in neural cells. **(d)** Expression of white matter microstructure phenotype-associated causal eGenes in the prenatal stage versus the postnatal stage in different cell types. For the locally estimated scatterplot smoothing (LOESS) plots, smooth curves are shown with 95% confidence intervals. P4, 13 PCW ≤ Age < 16 PCW; P8, Birth ≤ Age < 6 Months; P12, 12 ≤ Age < 20 Years; TS, tourette syndrome; ANX, anxiety disorder; MDD, major depressive disorder; ASD, autism spectrum disorder; BIP, bipolar disorder; SCZ, schizophrenia. Glia: glial cell, including microglia, astrocytes, oligodendrocytes, and oligodendrocyte precursor cells (OPCs). Neuro: neuronal cell, including inhibitory neurons and excitatory neurons.

For IDPs, we also analyzed the expression patterns of their causal eGenes in adult human brain cells (**Methods**). Consistent with our previous results of the causal eGenes predicted in IDPs (**Fig. 4b**), we observed a significantly elevated expression of the causal eGenes for white matter microstructure phenotypes in glial cells, particularly in astrocytes and microglia (**Fig. 5d**), which is expected given that the glial cells are the predominant constituents of white matter[104]. Additionally, we further explored the regional specificity of brain volume in different cell types using the causal eGenes, and our results showed that both subcortical and cortical regions exhibited stronger specificity to glial cells (**Supplementary Fig. 12**). These results suggest that glial cells, such as astrocytes and microglia, have a significant impact on the volume of brain regions in adults[105; 106].

### Association between IDPs and DBs based on the expression patterns of causal eGenes

The sophisticated functions of the brain are underpinned by its structural and regional connections and their intricate collaboration[107; 108]. However, the patterns of these connections at the cellular level remains elusive. To investigate the connection patterns within brain structures, we examined the relationships between regional brain volumes and white matter microstructure phenotypes, leveraging cell type-specific gene expression data from STAB2[109]. Our analysis showed that the regional brain volumes and white matter microstructure phenotypes tended to form distinct clusters (**Methods** and **Fig. 6a**). Additionally, at the regional level, we observed that cell type-specific gene expression can depict hemisphere specificity, where the same brain region was clustered into different modules across different cerebral hemispheres (*r* > 0.7, Spearman’s rank correlation). Furthermore, we performed similar analysis on DBs, and found that the expression levels of their causal eGenes showed concordance with the brain disorders and phenotypes (**Supplementary Fig. 13**). For example, education and intelligence were significantly positive associated with AD (*r* > 0.9, Spearman’s rank correlation), which demonstrates that high education and intelligence can reduce the risk of AD[110; 111].

**Fig. 6.**
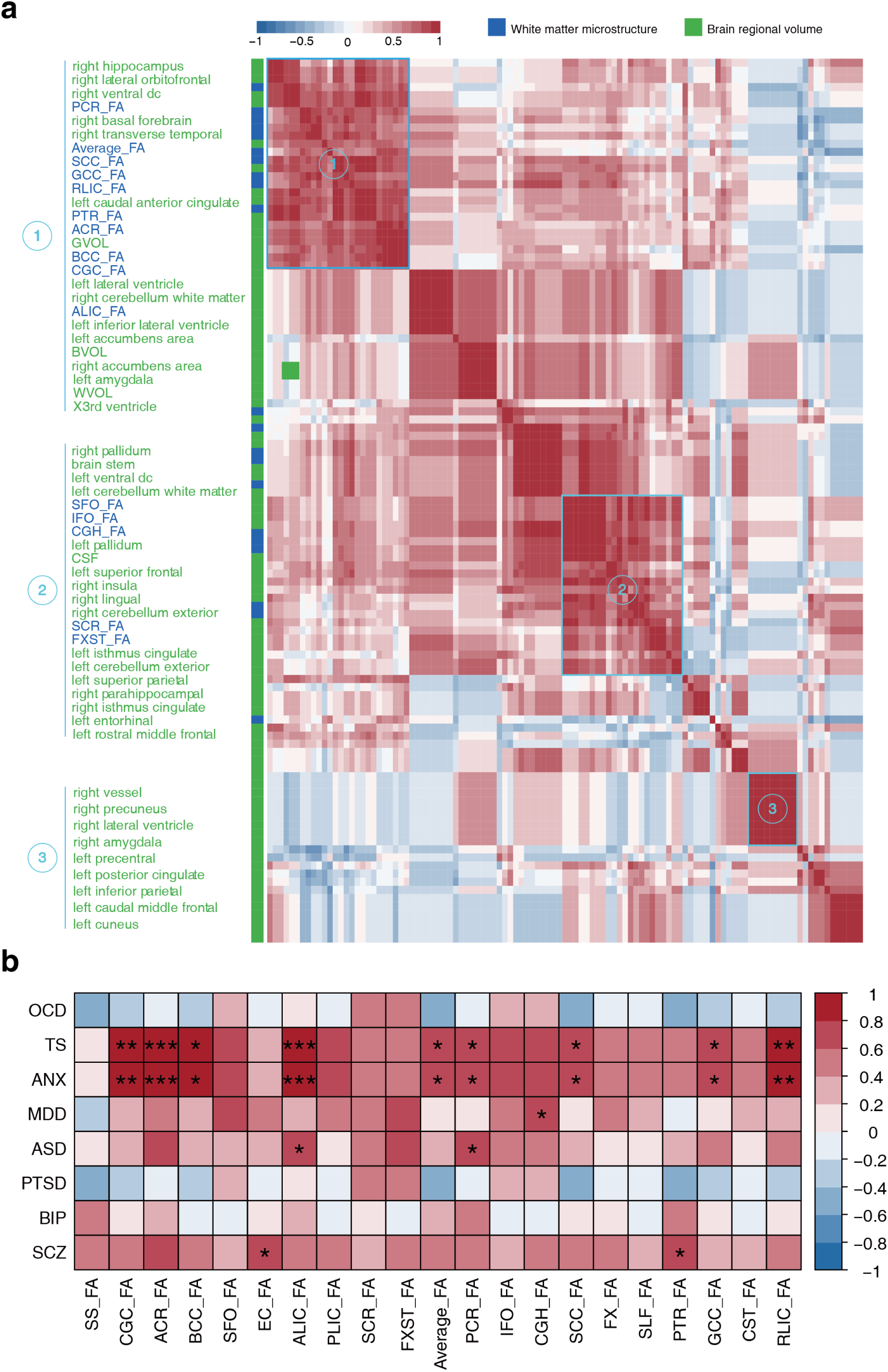
Biological associations within IDPs, and between white matter microstructure phenotypes and psychiatric disorders. **(a)** Cluster diagram within IDPs, evaluated by the expression of causal eGenes in eight cell types. Different colors represent different groups of IDPs. **(b)** Biological associations between white matter microstructure phenotypes and psychiatric disorders, evaluated by the expression of causal eGenes in eight cell types. Correlation values are based on Spearman’s correlation coefficient (**Methods**). *, FDR < 0.05; **, FDR < 0.01; ***, FDR < 0.001.

We have shown that the causal eGenes can mediate the associations between IDPs and DBs (**Fig. 4**). To investigate the biological plausibility of these causal associations, we explored the expression levels of the causal eGene in different cell types (**Methods**). Consistent with our previous findings on the putative causality routes between IDPs and DBs (**Fig. 4**), the FA of PTR and IFO was significantly associated with SCZ and GRT, respectively (**Fig. 6b** and **Supplementary Fig. 14**). Interestingly, we observed significant expression associations between several brain regional volumes and psychiatric disorders (FDR < 0.05, Spearman’s rank correlation test), including MDD and the left rostral-middle frontal cortex (FDR = 4.01×10^-45^, Spearman’s rank correlation test), and SCZ and the accumbens (FDR = 0.004, Spearman’s rank correlation test) (**Supplementary Fig. 15**).

## Discussion

In this study, we performed cell type-specific MR to dissect potential causal relationships between gene expression and brain-associated complex traits, leveraging several complementary methods to increase the result reliability. Our analysis identified 254 and 217 cell type-specific causal eGenes potentially causal for 112 IDPs and 26 DBs, respectively. Remarkably, the results exhibited high cell type specificity, with over 90% of causal eGene-IDP and 80% of causal eGene-DB associations being unique to a single cell type. The causal eGenes demonstrated widespread trait pleiotropy, particularly *DDHD2* in excitatory neurons, *XKR6* in inhibitory neurons, *MAPT* in astrocytes, and *ZSCAN31* in astrocytes. Primarily, we focused on the causal eGenes shared between brain-associated complex traits, and discovered that they were involved in biological processes such as memory and cognition, neurotransmitter regulation, protein binding, cell maintenance and signaling, and extracellular matrix dynamics. We also noted distinct patterns in brain cell types for these shared causal eGenes between the DB and IDP classes. According to the cell-specific spatiotemporal expression analysis, it indicated that expression of causal eGenes for PYSDs matched their developmental patterns, while causal eGenes for white matter microstructure phenotypes showed significantly higher expression in glial cells compared to neurons. Additionally, we found that the expression of causal eGenes of IDPs can establish connections within different brain regions, and confirm the association between the PTR and SCZ at the cell type level. These findings point to the genetic underpinnings at the cellular level in shaping brain structures, disorders and behaviors, as well as offer insights into the cellular mechanisms by which risk genes influence the relationships among them.

A significant overlap of cell type-specific causal eGenes was observed across different trait groups, and these genes were involved in biological processes such as memory and cognition, neurotransmitter regulation, protein binding, cell maintenance and cell signaling, and extracellular matrix dynamics. Substantial differences emerged in the types of brain cells for causal genes shared between IDPs and DBs; for instance, SCZ and the brain regional volume group shared genes across the broadest range of cell types, whereas neurodevelopmental disorders such as ASD, PTSD, and ADHD shared genes predominantly expressed in inhibitory neurons with brain structure groups. These findings point to the genetic underpinnings that operate at the cellular level in shaping brain structures, disorders and behaviors, and provide novel insights into the cellular mechanisms by which risk genes influence the relationships among them.

Previous studies have indicated an important but underrecognized role of excitatory neurons in AD etiology[112; 113]. In our research, six cell type-specific eGenes (*SYT14*, *MSH3*, *ICA1L*, *RGS14*, *C17orf97*, and *ZSCAN31*) that were causally associated with AD in excitatory neurons were identified (**Supplementary Table 5**). Among these genes, five (*MSH3*, *ICA1L*, *RGS14*, *C17orf97*, and *ZSCAN31*) demonstrated a consistent positive causal effect. Remarkably, several genes displayed specific associations with excitatory neurons or synaptic function. For instance, the *SYT14* gene is reportedly expressed in neural clusters, and is involved in calcium-dependent neurotransmitter release and synaptic vesicle trafficking[114; 115]; *ICA1L and RGS14* are implied in modulating synaptic signaling pathways in excitatory neurons[92; 116]; and *ZSCAN31* is a member of the zinc finger protein family that plays key roles in regulating neuronal excitability, which influences the mechanisms of intrinsic neural circuitry within the excitatory neuron subtype[117; 118]. This consistency implies that these genes may influence AD progression through common and cumulative mechanisms that affect neuronal functions. Additionally, our findings revealed significant overlaps between AD and brain structural groups with respect to cell type-specific causal eGenes in excitatory neurons (**Fig. 3**), including *ZSCAN31, SYT14* and *ICA1L* (**Supplementary Figs. 6 and 7**). These results collectively enhance our understanding of the mechanistic links among genes, excitatory neurons, AD, and brain structure.

The *ZSCAN31* gene is one of the genes that shows the most pervasive trait pleiotropy. Previously, a study revealed that *ZSCAN31* was significantly downregulated in the hippocampus and frontal cortex of SCZ patients[98]. This study lends credence to our findings that the *ZSCAN31* gene in excitatory neurons has a negative causal effect on the volume of the right hippocampus and WVol (**Supplementary Table S4**). Differentially, across the 11 traits from the four groups with which the *ZSCAN31* gene in astrocytes was causally associated, including MDD and SCZ from the psychiatric disorder group, GRT from the behavioral-cognitive phenotype group, right supramarginal volume from the brain regional volume group, and 7 white matter microstructure phenotypes (the mean FA of GCC, PTR, SLF, SCC, BCC, ACR, and average FA across all tracts), all demonstrated a positive causal effect (**Fig. 3b** and **Supplementary Figs. 6-7**). This divergence in the causal effect direction of *ZSCAN31* across different cell types indicates cell type-specific regulation of the gene to brain-associated complex traits, further refining the discovery of this literature[98]. Based on these results, we further deduced three coherent routes involving *ZSCAN31* in astrocytes, IDPs and DBs (**Fig. 4** and **Supplementary Table S8**), including *ZSCAN31* in astrocytes – SCZ – PTR, which indicates that *ZSCAN31* expression in astrocytes may contribute to the causal effect of SCZ disease progression on PTR microstructural changes. Consistently, leveraging external single-cell data suggested that the causal eGenes in different cell types mediate the causal associations between SCZ and PTR (**Fig. 6b**), shedding light on the cell type-specific eGene regulation of brain-associated complex traits.

Our study offers new insights into the genetic underpinnings shared between risky behaviors and brain regions related to executive and visual functions at the cellular level. Initially, we discovered significant sharing of causal eGenes within inhibitory neurons between risky behaviors (e.g., GRT and ASP) and the brain regional volume and white matter microstructure phenotype groups (**Fig. 3a**). Further exploration revealed that the individual brain regions that significantly shared causal eGenes related to risky behaviors were primarily related to executive and visual functions, including the FXST, IFO, UNC, corona radiata, orbitofrontal cortex, isthmus cingulate, and ventral diencephalon (**Supplementary Fig. 5**). Upon looking at the individual eGene, we discovered that the *XKR6* gene was shared between risky behaviors (including ASP and GRT) and executive/visual-related IDPs (including the volume of right lateral orbitofrontal cortex, left isthmus cingulate, and left/right ventral diencephalon; the mean FA of FXST, IFO, and UNC). Finally, using the external expression data of the causal eGenes across different cell types, we discovered a significant correlation between GRT and IFO, as well as between GRT and FXST (**Fig. 6b**), which suggests the mediating role of cell type-specific causal eGenes in the associations between them. Previously, studies have highlighted the presence of region-specific inhibitory neuron types in the primary visual cortex[119], the influence of region-specific inhibitory activity of neurons on risky behaviors[64], and the association of variants within the *XKR6* gene with risky behaviors[86]. These pairwise evidences collectively strengthen the credibility of our findings and highlight the need for future investigation into the genetic and cellular underpinnings linking risky behaviors to relevant brain regions.

We believe that multiple future directions can be explored based on our work. First, the lower number of *cis*-eQTLs in endothelial cells and pericytes, compared to other cell types (**Supplementary Table 3**)[10], resulted in fewer identified causal genes within these cell types (**Fig. 1b**). Nevertheless, prior research has suggested the important role of these cell types in the pathogenesis of brain disorders[120–122]. With a larger amount of brain single-cell-derived gene expression data, we are more likely to identify more meaningful genes involved in brain illnesses within these cell types. Second, when performing MR between IDPs and DBs, half of the GWASs of DBs were removed due to the inclusion of samples from the popular dataset UK Biobank (UKB)[123], which reduced the completeness of the results when characterizing putative causality routes among eGenes, IDPs and DBs (**Fig. 4**). Future work could strive to collect as many mutually independent GWAS datasets of brain-associated complex traits as possible to gain a more comprehensive understanding of the putative causality routes connecting eGenes, IDPs and DBs. Third, while our research has highlighted possible causal relationships, these findings do not confirm exact causality and need to be verified by randomized controlled trials, the gold standard for causal inference. There is still a long road lays ahead in early treatment and therapeutic development, and thus careful interpretation of the results is needed.

To summarize, our study provides a systematic investigation of causal genes in the cell type context for brain-associated complex traits, as well as evidence of shared genetic foundations operating at the cellular level between brain structure and disorders/behaviors. We hope that the connections discovered among genes, cell types and complex traits will serve as starting points for the development of valuable therapeutic targets for brain structural abnormalities, disorders and behaviors.

## Methods

### Cell type-specific *cis*-eQTL statistics

We obtained the recently published cell type-specific *cis*-eQTL summary statistics dataset[10], which includes effect sizes for a total of 67,815,924 SNP-gene pairs across 4,690,822 SNPs and 17,881 gene transcripts. The data covers eight major brain cell types, including astrocytes, excitatory neurons, inhibitory neurons, oligodendrocytes, oligodendrocyte precursor cells (OPCs), microglia, endothelial cells, and pericytes (**Fig. 1**). The eQTL analysis was conducted using high-quality genotype and single-nuclei RNA sequencing data from 192 individuals derived from the prefrontal cortex, temporal cortex, and white matter. All individuals were within three standard deviations of the mean for the first and second principal components of the European-ancestry populations from the 1000 Genomes Project[10].

### GWAS summary statistics for brain disorders and behaviors

We obtained publicly available GWAS summary statistics for 26 brain disorders and behaviors (DBs), including eight behavioral-cognitive phenotypes, eight neurological disorders, and 10 psychiatric disorders (**Supplementary Table 1**). Most of these GWAS summary statistics were published recently and based on meta-analyses with a large sample size, with an average sample size of over 280,000 individuals across these datasets. All of these GWAS summary statistics were primarily of European ancestry. There are no overlapping individuals between these GWAS datasets and cell type-specific *cis*-eQTL summary statistics.

The sample size for each of the GWAS summary statistics datasets was listed as follows:

- Behavioral-cognitive phenotypes

(1) Automobile speeding propensity (ASP): 404,291 individuals[86]
(2) Childhood intelligence (CI): 12,441 individuals[124]
(3) Drinks per week (DPW): 941,280 individuals[125]
(4) Educational attainment (EA): 766,345 individuals[126]
(5) General risk tolerance (GRT): 466,571 individuals[86]
(6) Intelligence (INT): 269,867 individuals[127]
(7) Neuroticism (NEU): 390,278 individuals[128]
(8) Smoking cessation (SC): 547,219 individuals[125]
- Neurological disorders

(1) Alzheimer’s disease (AD): 71,880 cases and 383,378 controls[129]
(2) Amyotrophic lateral sclerosis (ALS): 20,806 cases and 59,804 controls[130]
(3) Epilepsy (EPI): 15,212 cases and 29,677 controls[131]
(4) Intracerebral hemorrhage (ICH): 1,545 cases and 1,481 controls[132]
(5) Insomnia (INS): 109,548 cases and 277,440 controls[133]
(6) Ischemic stroke (IS): 67,162 cases and 454,450 controls[134]
(7) Multiple sclerosis (MS): 47,429 cases and 68,374 controls[135]
(8) Parkinson’s disease (PD): 33,674 cases and 449,056 controls[136]
- Psychiatric disorders

(1) Attention-deficit/hyperactivity disorder (ADHD): 20,183 cases and 35,191 controls[137]
(2) Anxiety disorder (ANX): 25,453 cases and 58,113 controls[138]
(3) Autism spectrum disorder (ASD): 18,381 cases and 27,969 controls[139]
(4) Alcohol use disorder (AUD): 11,569 cases and 34,999 controls[140]
(5) Bipolar disorder (BIP): 41,917 cases and 371,549 controls[141]
(6) Major depressive disorder (MDD): 135,458 cases and 344,901 controls[142]
(7) Obsessive-compulsive disorder (OCD): 2,688 cases and 7,037 controls[143]
(8) Post-traumatic stress disorder (PTSD): 9,831 cases and 19,225 controls[144]
(9) Schizophrenia (SCZ): 76,755 cases and 243,649 controls[145]
(10) Tourette syndrome (TS): 4,819 cases and 9,488 controls[146]

### GWAS summary statistics for imaging-derived phenotypes

We obtained publicly available GWAS summary statistics for 123 brain imaging-derived phenotypes (IDPs), including 101 brain regional (and total) volume phenotypes and 22 white matter microstructure phenotypes (**Supplementary Table 2**). The GWAS summary statistics for brain volume phenotypes were processed by Zhao et al.[147], with a sample size of 19,629 participants of European ancestry from UKB. The brain volumes include total brain volume, gray matter, white matter and cerebrospinal fluid, and were labeled by Mindboggle-101 atlases[148]. The GWAS summary statistics for white matter tracts were processed by Zhao et al.[18], with a sample size of 33,292 participants of European ancestry from UKB. The white matter tracts were segmented by the JHU ICBM-DTI-81 white-matter atlas[149–151], and the microstructure of the above-mentioned tracts is measured by the mean fractional anisotropy (FA).

### Transcriptome-wide association study

To investigate the potential links between gene expression levels and these brain-associated complex traits, we conducted a transcriptome-wide association study (TWAS) for the GWAS summary statistics of 123 IDPs and 26 DBs[26]. The prediction weights and covariance used for TWAS analysis were obtained from two resources: (1) PredictDB Data Repository (https://predictdb.org/), where the prediction models were built based on the GTEx V8 expression data[152] from 13 brain regions including Amygdala, Anterior Cingulate Cortex (BA24), Caudate (basal ganglia), Cerebellar Hemisphere, Cerebellum, Cortex, Frontal Cortex (BA9), Hippocampus, Hypothalamus, Nucleus accumbens (basal ganglia), Putamen (basal ganglia), Spina cord (cervical c-1) and Substantia nigra; (2) CMC-derived DLPFC prediction models (https://github.com/laurahuckins/CMC_DLPFC_prediXcan), which were built based on the CommonMind Consortium expression data[153] from dorsolateral pre-frontal cortex (DLPFC) tissue. The TWAS analysis was implemented using MetaXcan[26]. The genes that were potentially associated with the brain-associated complex traits were retained for subsequent MR analysis. Accordingly, the gene-trait pairs reaching a nominally significant threshold (*P* < 0.05) in at least one brain tissue were preserved with their suggestive evidence of association.

### Primary Mendelian randomization analysis

To investigate the causal effects of cell type-specific gene expression on brain-associated complex traits, we performed two-sample MR analysis using four complementary MR methods: two single-SNP methods, including SMR[4] and Wald radio[27], and two multi-SNP methods, including PMR-Egger[6] and GSMR[28]. Details on the implementation of each MR method are provided as follows (**Supplementary Fig. 1**).

1. The SMR method tests whether the expression level of a gene is associated with a complex trait under the assumption of either causality or pleiotropy[4]. It is specifically designed for gene expression as the exposure, and requires only one SNP as the instrument. For the instrument selection, the top *cis*-eQTL associated with each gene expression in a given cell type at genome-wide significance was selected (*P* < 5×10^-8^). The SMR incorporates the HEIDI test to distinguish whether the presence of linkage influences the association result[4]. If the *P*-value of the HEIDI test is below 0.05, it indicates the presence of linkage, and the result is subsequently discarded. Both the SMR method and HEIDI test were implemented using the SMR software (version 1.3.1, https://yanglab.westlake.edu.cn/software/smr).
2. The Wald ratio method computes the change in disease risk per standard deviation change in gene expression, explained through the instrumental *cis*-eQTL for that gene. It is a universal method applicable to any kind of exposure, and requires only one SNP as the instrument. For the instrument selection, LD clumping (r^2^ < 0.1, window = 250 kb, 1000 Genomes EUR reference panel) was first performed for the *cis*-eQTLs associated with each gene expression at genome-wide significance (*P* < 5×10^-8^) using PLINK v1.90[154]. Subsequently, the independent *cis*-eQTL(s) for each gene expression in the given cell type were selected as the instrument(s). After instrument selection, 86.6% of cell type-specific genes were associated with one valid instrumental *cis*-eQTL, thus Wald ratio can be applied in most analyses. For the remaining 13.4% of cell type-specific genes with more than one valid instrument, the inverse-variance weighted (IVW, fixed-effects) method was implemented as a supplementary method[155], which is frequently paired with Wald ratio in relevant literature[25; 156]. Both the Wald ratio and IVW methods were implemented using the R package TwoSampleMR (version 0.5.10)[157]. Additionally, the *harmonise_data()* function in TwoSampleMR was used to harmonize the effect alleles and SNP effects between the exposure and outcome. The heterogeneity test (function ‘*mr_heterogeneity*’, *P* < 0.05) and pleiotropy test (function ‘*mr_pleiotropy_test*’, *P* < 0.05) were conducted to detect the presence of heterogeneity and horizontal pleiotropy in IVW results.
3. The PMR-Egger method estimates the causal effect of the expression level of a gene on the trait in the presence of horizontal pleiotropy within a MR likelihood framework[6]. It is specifically designed for gene expression as the exposure. It can effectively handle multiple correlated variants as instruments as well as account for the linkage disequilibrium (LD) between the variants. For the instrument selection, the *cis*-eQTLs (clumping *r^2^* < 0.9, window = 250 kb, 1000 Genomes EUR reference panel) associated with each gene expression in a given cell type at genome-wide significance (*P* < 5×10^-8^) were selected. The PMR-Egger method was implemented using the R package PMR (version 1.0). The LD correlation matrix for instruments used in PMR-Egger analysis were derived from 1000 Genomes EUR reference panel, and the maximum iteration in the algorithm was set at 5000.
4. The GSMR method is an extension of the SMR method, which estimates the causal effect of an exposure on an outcome, leveraging power from multiple variants while accounting for LD among them[28]. It is a universal method applicable to any kind of exposure. For the instrument selection, the *cis*-eQTLs (clumping *r^2^* < 0.9, 1000 Genomes EUR reference panel) associated with each gene expression in a given cell type at genome-wide significance (*P* < 5×10^-8^) were selected. The GSMR analysis incorporates the HEIDI-outlier test to identify and exclude instruments exhibiting significant pleiotropic effects on the outcome, using a *P*-value threshold of 0.05. Both the GSMR method and HEIDI-outlier test were implemented using the R package gsmr (version 1.1.0).

A gene is deemed putatively causal for a trait within a given cell type (called putatively causal cell type-specific eGene or cell type-specific causal eGene for short) if the results satisfy the following requirements: (1) significant in at least one significant single-SNP method after false discovery rate (FDR) correction [Controlling the False Discovery Rate: of practical and powerful approach two multiple testing] (corrected *P* < 0.05); (2) significant in at least one significant multi-SNP method after FDR correction (corrected *P* < 0.05); (3) exhibiting consistent causal effect directions across all four methods, thus less likely to be a false positive.

Furthermore, we calculated the *F*-statistic for each instrument using the Cragg-Donald statistic to access instrument strength[158]: 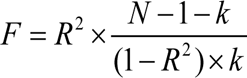, where 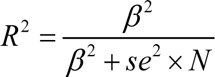 represents the proportion of exposure variance explained by the instrument, *N* is the sample size of the instrument from the eQTL summary statistics, *β* is the effect size of the instrument from the eQTL summary statistics, *se* is the standard error of the effect size, and *k* is the number of instruments used in the MR estimate (*k* = 1 for a single instrument). The results in which the averaged *F*-statistic across all instruments for that gene exceeded 10 were retained.

### Significance of causal eGenes sharing

To test whether cell type-specific causal eGenes significantly shared between two brain-associated complex traits, the hypergeometric test was applied. The background gene set was derived by multiplying the number of cell types by the total number of protein-coding genes, lincRNA, and processed transcripts (the main three biotypes of the identified cell type-specific causal eGenes showed in **Supplementary Tables 4** and **5**) in the Ensemble database GRCh37 (release 87) coordinate file (8 × 26,861). Furthermore, to test the sharing of cell type-specific causal eGenes within a particular cell type, the hypergeometric test with the background gene set being the total number of protein-coding genes, lincRNA and processed transcripts from the GRCh37 (release 87) coordinate file (26,861) was used. The FDR significance threshold was empirically set at 0.05 to select the significant cell types.

### Gene set enrichment analysis

To elucidate biological processes implicated in different groups of brain-associated complex traits, we further looked at gene sets enriched among cell type-specific causal eGenes shared within each trait group (shared by at least two traits within each group) and between trait groups (shared by at least one trait in one group and at least one in another). We utilized all gene sets from the Biological Process (BP), Molecular Function (MF), and Cellular Component (CC) subontologies in the Gene Ontology (GO) database for this enrichment analysis[159; 160]. The GO enrichment analysis was implemented using the function *enrichGO()* in R package clusterProfiler (version 4.10.0)[46].

### Replication of gene-trait associations

Replication was accessed for the identified causal association between eGenes and brain-associated complex traits in our MR analysis. We performed replication using data from three external resources: (1) We obtained *cis*-eQTL summary statistics from the BrainMeta portal (https://yanglab.westlake.edu.cn/data/brainmeta/cis_eqtl/), which was derived using RNA sequencing data of 2,865 brain cortex samples from 2,443 unrelated individuals of European ancestry[29]. The same MR analysis procedure was applied to this data to infer the causal relationships between cortical gene expression and 149 brain-associated complex traits. An eGene-trait pair was deemed replicated if it achieved nominal significance in at least one MR method within the BrainMeta data. (2) We extracted differentially expressed genes and transcripts (at 5% FDR) in ASD, SCZ and BIP from PsychENCODE[23], which leveraged in vivo gene expression profiles to pinpoint specific alterations in transcriptome-wide isoform levels across the three major psychiatric disorders. (3) We obtained previously documented associations between genes and complex traits from the NHGRI-EBI GWAS Catalog (version 2023-06-09, www.ebi.ac.uk/gwas/)[34]. For each of these external resources, we evaluated the replication of eGene-trait pairs by calculating the replication rate (number of replicated eGene-trait pairs / number of both replicated and not replicated eGene-trait pairs) and the *P*-value through the hypergeometric test. Replication was assessed only for traits that appeared in both the discovery and replication studies.

### Mendelian randomization between brain IDPs and DBs

In order to characterize putative causality routes among cell type-specific genes, IDPs and DBs, we extended to perform bidirectional MR analysis for inferring the relationships between the above DBs and IDPs. Since the GWAS of IDPs was conducted in participants from the UKB, to minimize the issue of sample overlap in MR, we utilized the GWAS of DBs that did not include UKB participants. Then 13 GWAS of DBs (including CI, EPI, ICH, IS, ALS, MS, OCD, TS, ADHD, AUD, ASD, PTSD, and SCZ) were retained for downstream analysis. Differing from MR analysis focused on gene expression as the exposure, which typically has a single independent genome-wide significant eQTL instrument, it often involves multiple independent genome-wide significant independent instruments for DBs or IDPs as the exposure. Therefore, we primarily employed multi-SNP MR methods here, which include PMR-Egger, GSMR and methods in R package TwoSampleMR (IVW[155], MR-Egger[161], Weighted median[162], Simple mode[163], Weighted mode[164]). For the instrument selection, it was selected as variants (clumping *r^2^* < 0.6) associated with each exposure at genome-wide significance (*P* < 5×10^-8^) in PMR-Egger; variants (clumping *r^2^*< 0.1) associated with each exposure at genome-wide significance (*P* < 5×10^-8^) in GSMR; and variants (clumping *r^2^* < 0.1) for each exposure at genome-wide significance (*P* < 5×10^-8^) in methods in R package TwoSampleMR. The heterogeneity test (function ‘*mr_heterogeneity*’, *P* < 0.05) and pleiotropy test (function ‘*mr_pleiotropy_test*’, *P* < 0.05) were conducted to detect the presence of heterogeneity and horizontal pleiotropy in TwoSampleMR results. A trait is deemed putatively causal for another trait if the results satisfy the following requirements: (1) significant in at least two MR methods after FDR correction (corrected *P* < 0.05); (2) exhibiting consistent causal effect directions across all methods. Additionally, IDP-DB pairs showing bidirectional causality were excluded from the results to ensure clarity in determining directional influences.

### Analysis of expression patterns of causal eGenes using external single-cell data

External normalized single-cell gene expression data were sourced from our prior study, STAB2[109]. We extracted the cell expression data of causal eGenes associated with IDPs and DBs from the eight cell types used in this work (i.e., microglia, astrocytes, oligodendrocytes, OPCs, inhibitory neurons, excitatory neurons, endothelial cells, and pericytes) from STAB2. To assess the relationship between IDPs and DBs at the cell type level, we first collated expression data of the causal eGenes for each IDP or DB within the specified cell types. We then created an eight-dimensional vector for each IDP or DB, with each value representing the mean expression value of the relevant causal eGenes in a cell type, facilitating a direct comparison. Finally, Spearman’s rank correlation coefficient was used to evaluate the associations between IDPs and DBs based on the cell type-specific expression of the corresponding causal eGenes. Additionally, hierarchical clustering (complete linkage) was employed to investigate the internal relationships within IDPs and DBs, leveraging these correlation coefficients.

## Data Availability

All data produced in the present work are contained in the manuscript

