## Supplementary Figures for "Deciphering causal relationships between cell type-specific genetic factors and brain imaging-derived phenotypes and disorders"

**Deciphering cell type-specific causal genetic effects on brain imaging-derived phenotypes and disorders using single-cell Mendelian randomization**

Anyi Yang, Xingzhong Zhao, Yucheng T. Yang, Xing-Ming Zhao

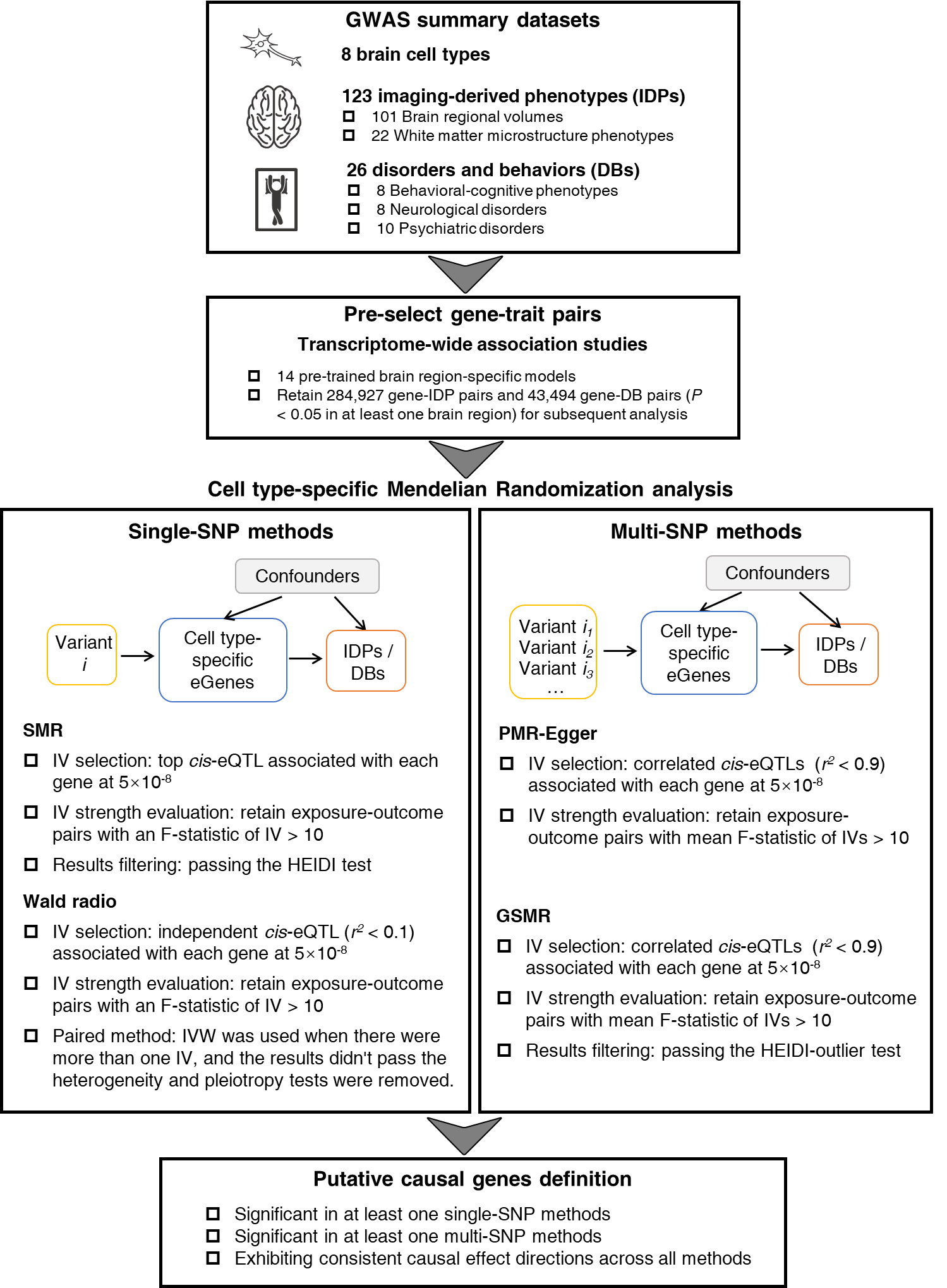

**Supplementary Fig. 1. Study workflow for the primary Mendelian randomization analysis**

IV: instrumental variables.

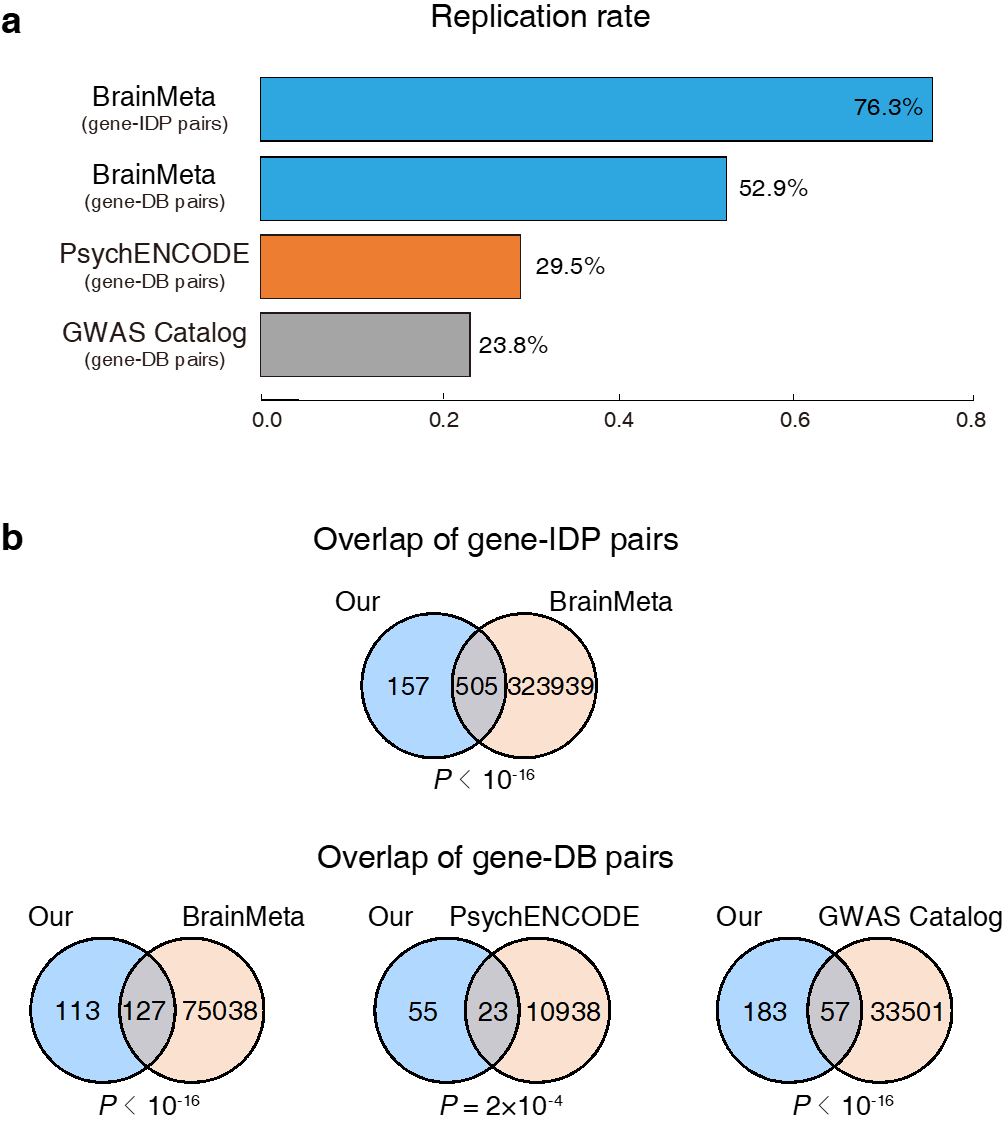

**Supplementary Fig. 2. Replication results for the identified association between causal eGenes and brain-associated complex traits**

We assessed the reproducibility of eGene-IDP pairs using cortical *cis*-eQTL summary statistics from BrainMeta^1^. We assessed the reproducibility of eGene-DB pairs using three distinct datasets: the cortical *cis*-eQTL summary statistics from BrainMeta^1^, differentially expressed genes and transcripts from PsychENCODE^2^, and documented associations between genes and complex traits from GWAS Catalog^3^. **(a)** Replication rates for predicted eGene-IDP pairs in BrainMeta, and for predicted eGene-DB pairs across BrainMeta, PsychENCODE and GWAS Catalog. **(b)** Overlap of gene-IDP pairs and gene-DB pairs between our study and BrainMeta, PsychENCODE and GWAS Catalog, assessed using the hypergeometric test.

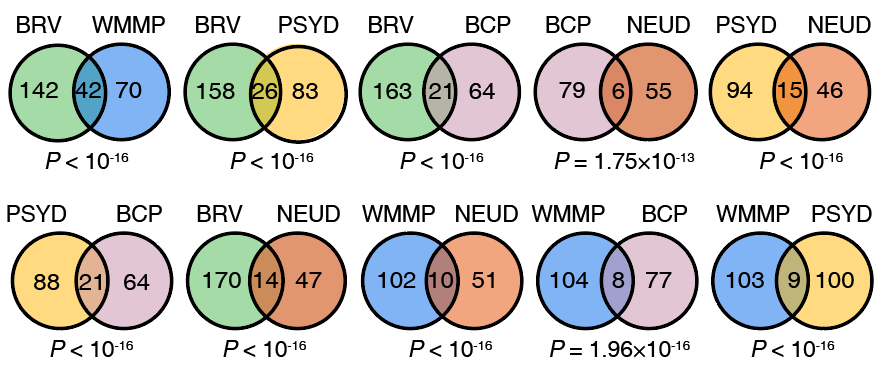

**Supplementary Fig. 3. Overlap of causal eGenes between each pair of trait groups**

The overlaps were assessed using the hypergeometric test to determine statistical significance. BRV: Brain regional volume; WMMP: White matter microstructure phenotype; BCP: Behavioral-cognitive phenotype; NEUD: Neurological disorder; PSYD: Psychiatric disorder.

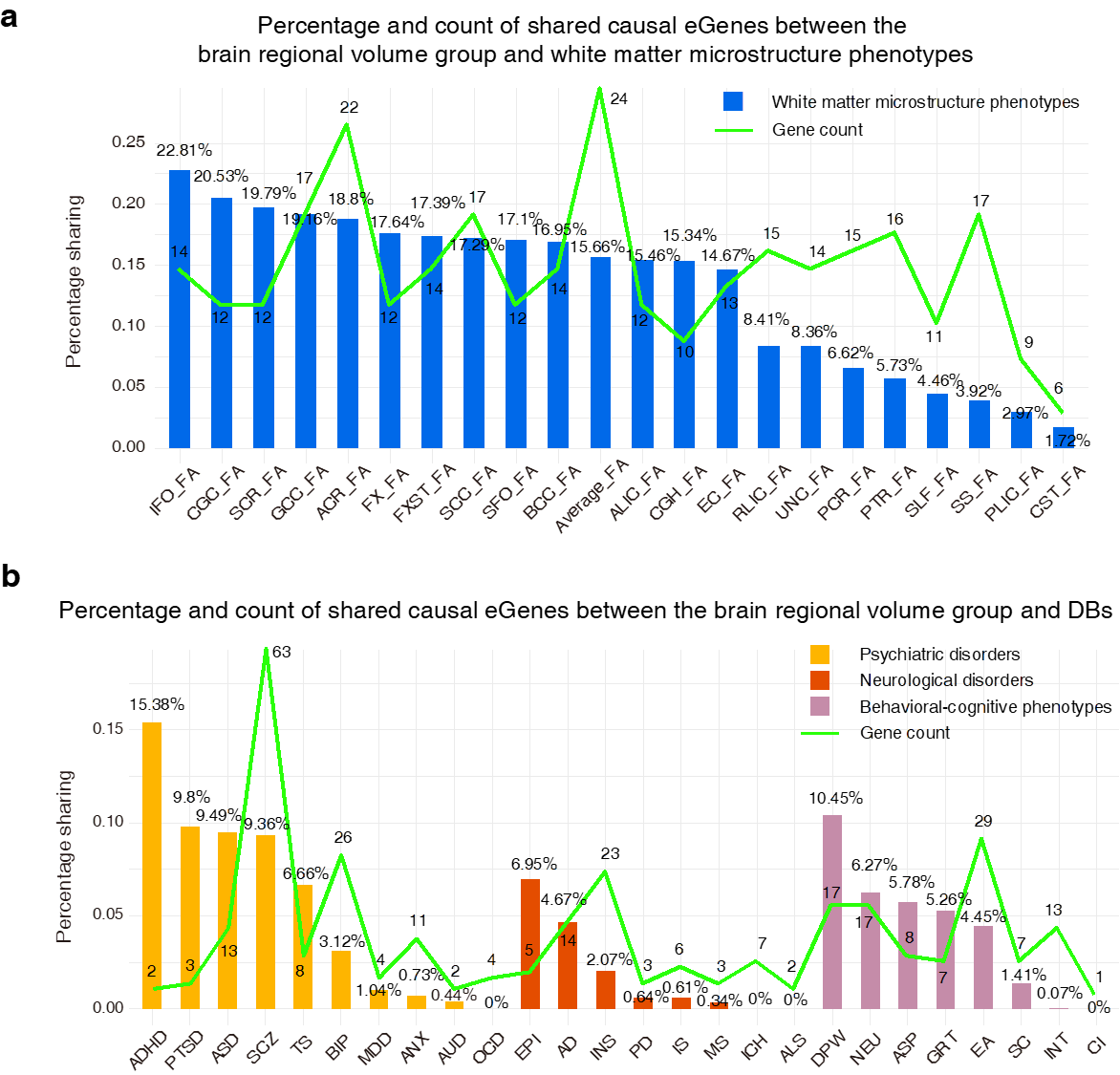

**Supplementary Fig. 4. Percentage and count of shared cell type-specific causal eGenes between different trait types**

**(a)** Percentage and count of shared cell type-specific causal eGenes between the brain regional volume group and white matter microstructure phenotypes. **(b)** Percentage and count of shared cell type-specific causal eGenes between the brain regional volume group and DBs.

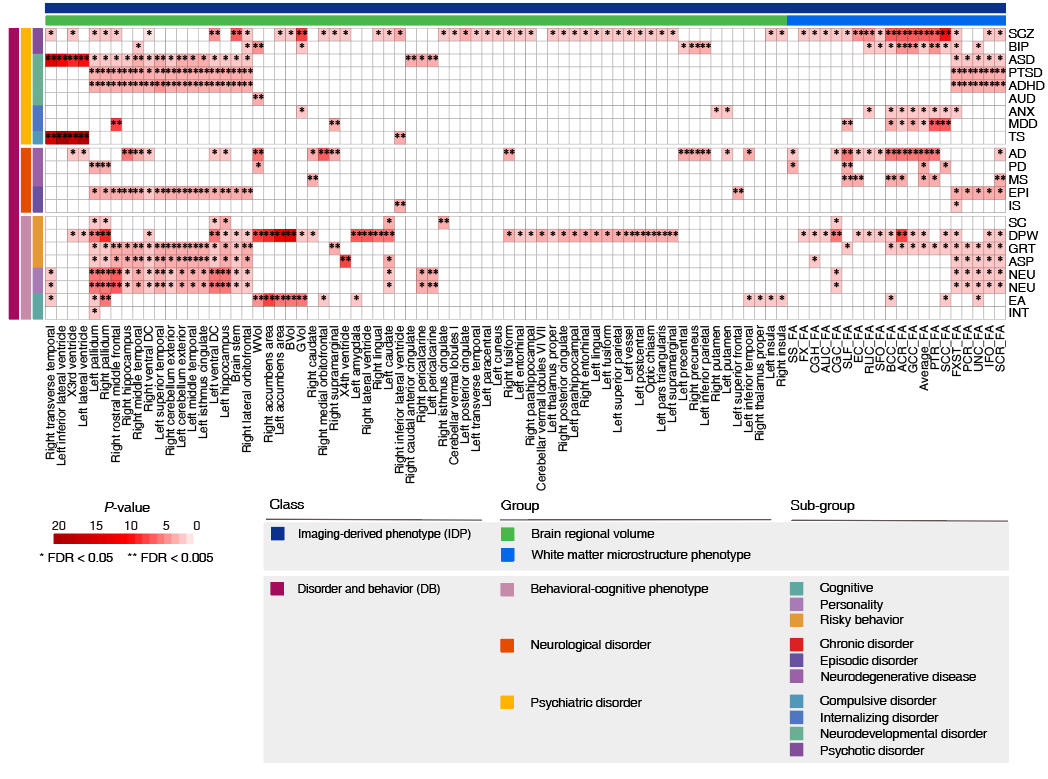

**Supplementary Fig. 5. Heatmap of shared causal cell type-specific eGenes between IDPs and DBs**

The overlaps of causal cell type-specific eGenes between two types of traits were assessed using the hypergeometric test to determine statistical significance.

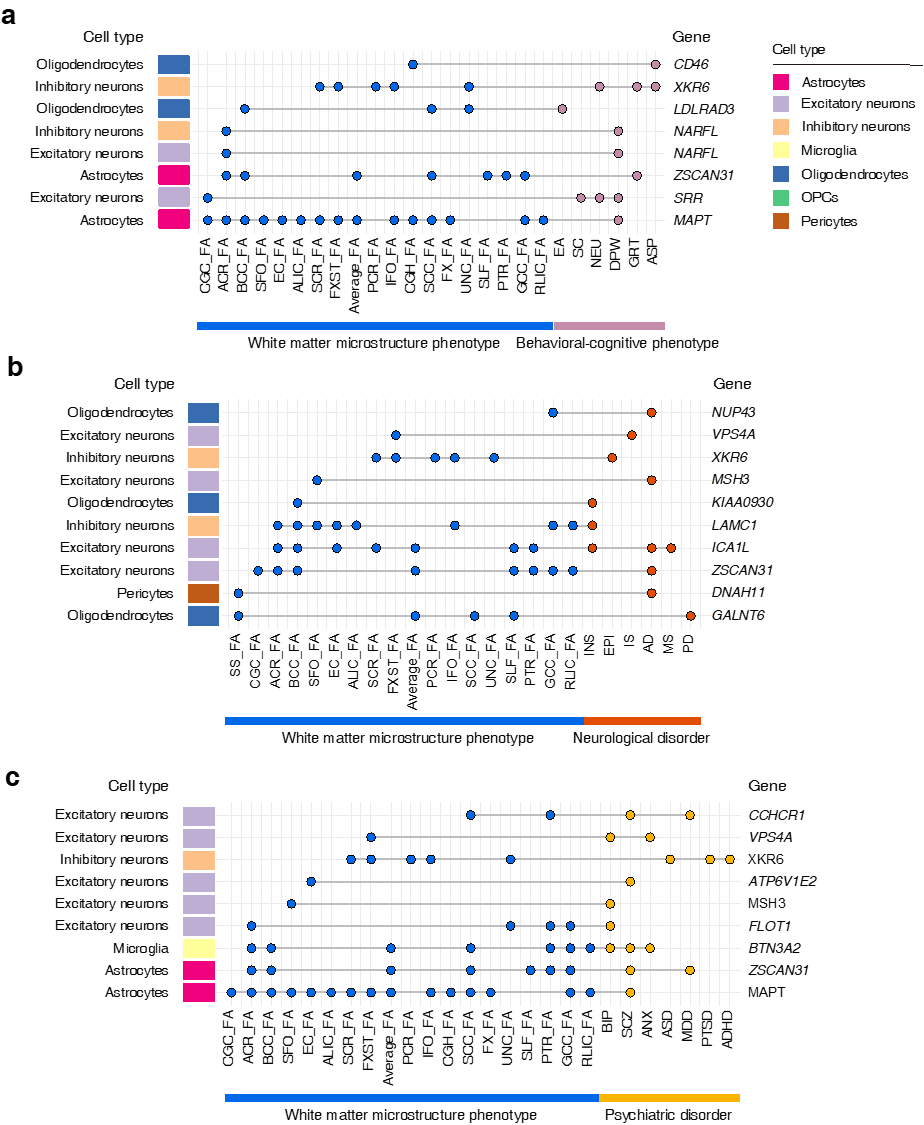

**Supplementary Fig. 6. Cell type-specific causal eGenes shared between white matter microstructure phenotypes and behavioral-cognitive phenotypes (a), neurological disorders (b), and psychiatric disorders (c).**

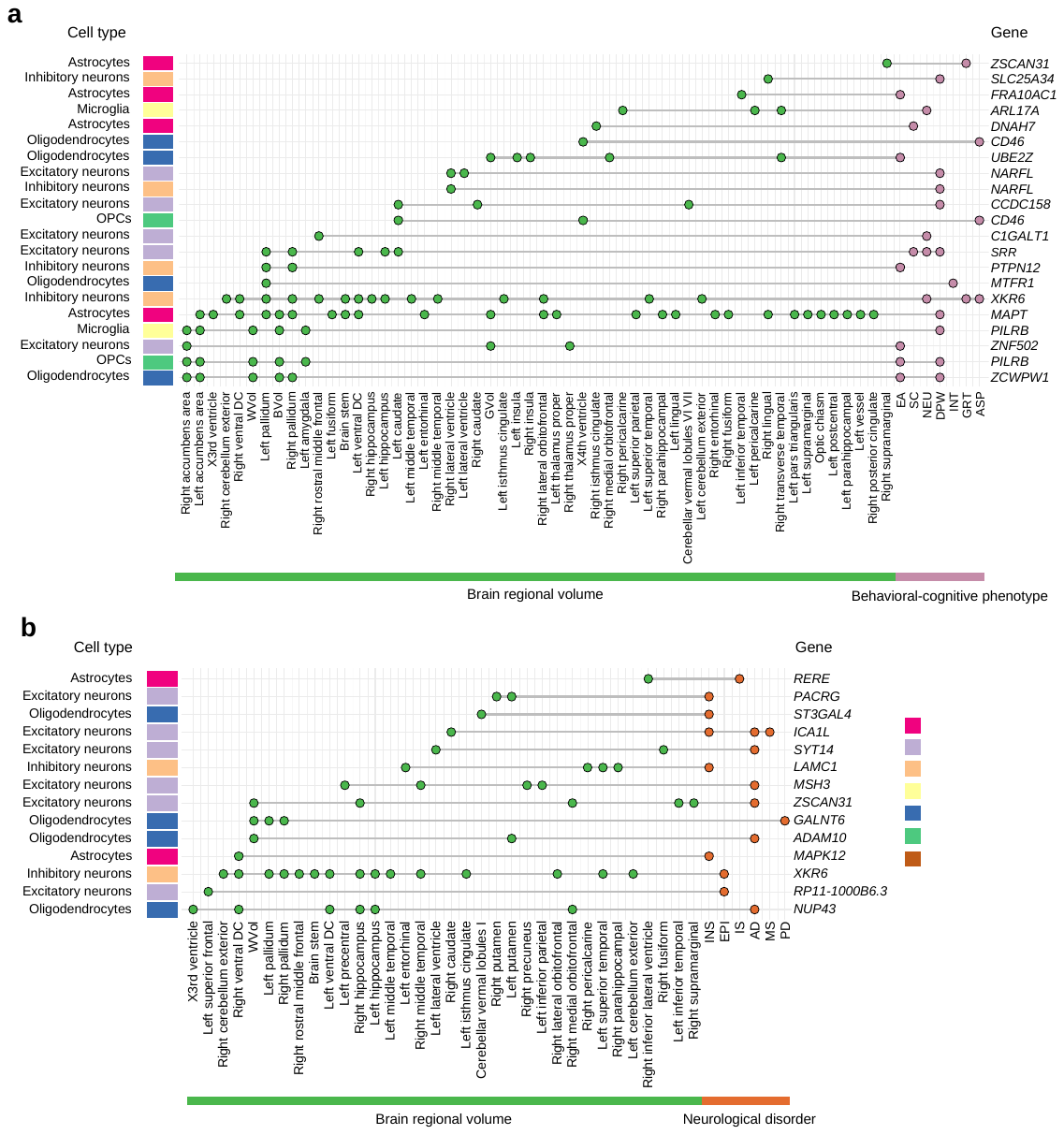

**Supplementary Fig. 7. Cell type-specific causal eGenes shared between brain regional volumes and behavioral-cognitive phenotypes (a), as well as neurological disorders (b).**

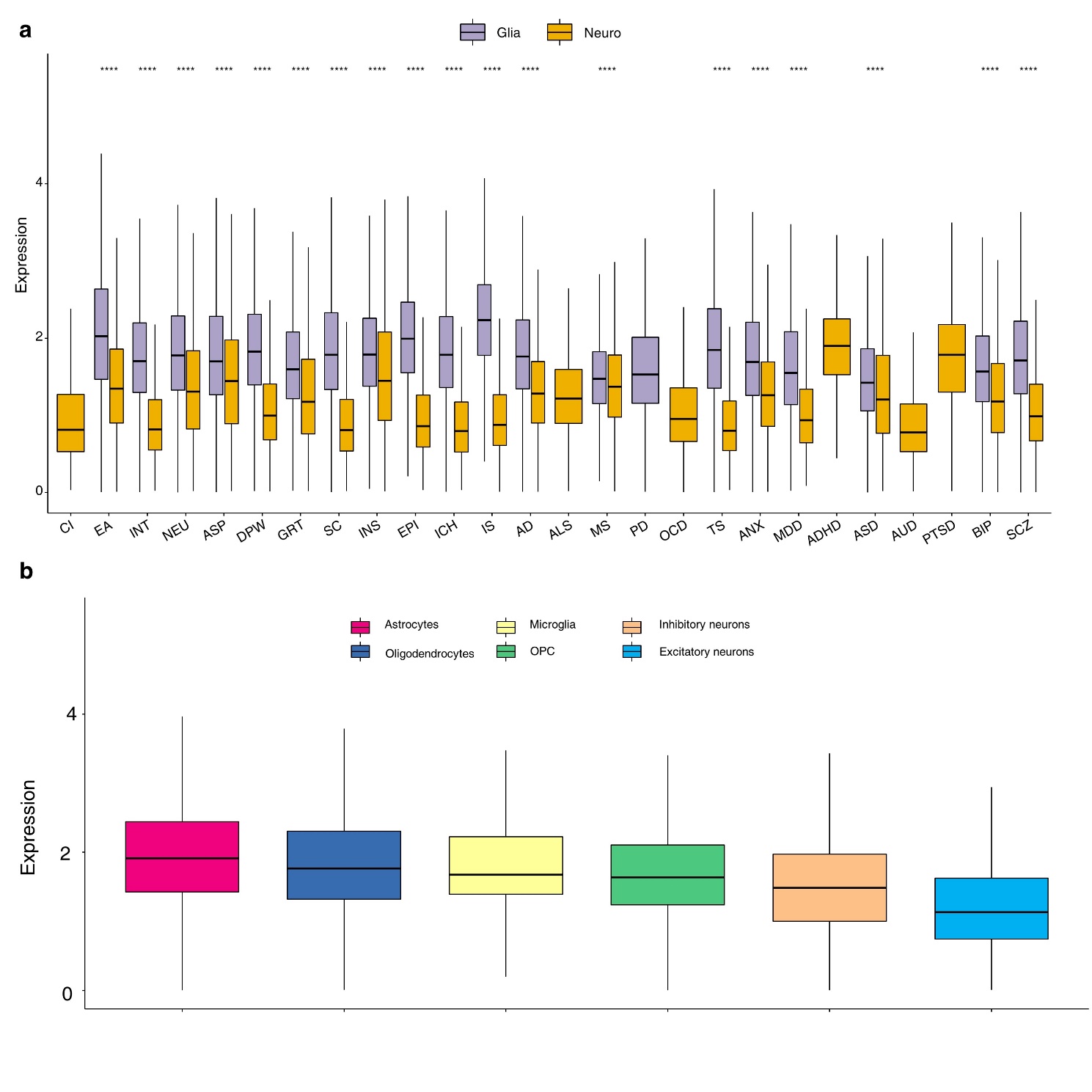

**Supplementary Fig. 8.** **Expression levels of the causal eGenes of 26 DBs from glial and neural cells**

**(a)** Expression differences in glial and neural cell types for various DBs, with significance indicated as ***, *P* < 0.001. **(b)** Detailed expression profiles for causal eGenes of 26 DBs in glial and neural cell types. Glia: glial cells, including astrocytes, microglia, oligodendrocytes, and oligodendrocyte precursor cells (OPCs). Neuro: neuronal cells, including inhibitory neurons and excitatory neurons.

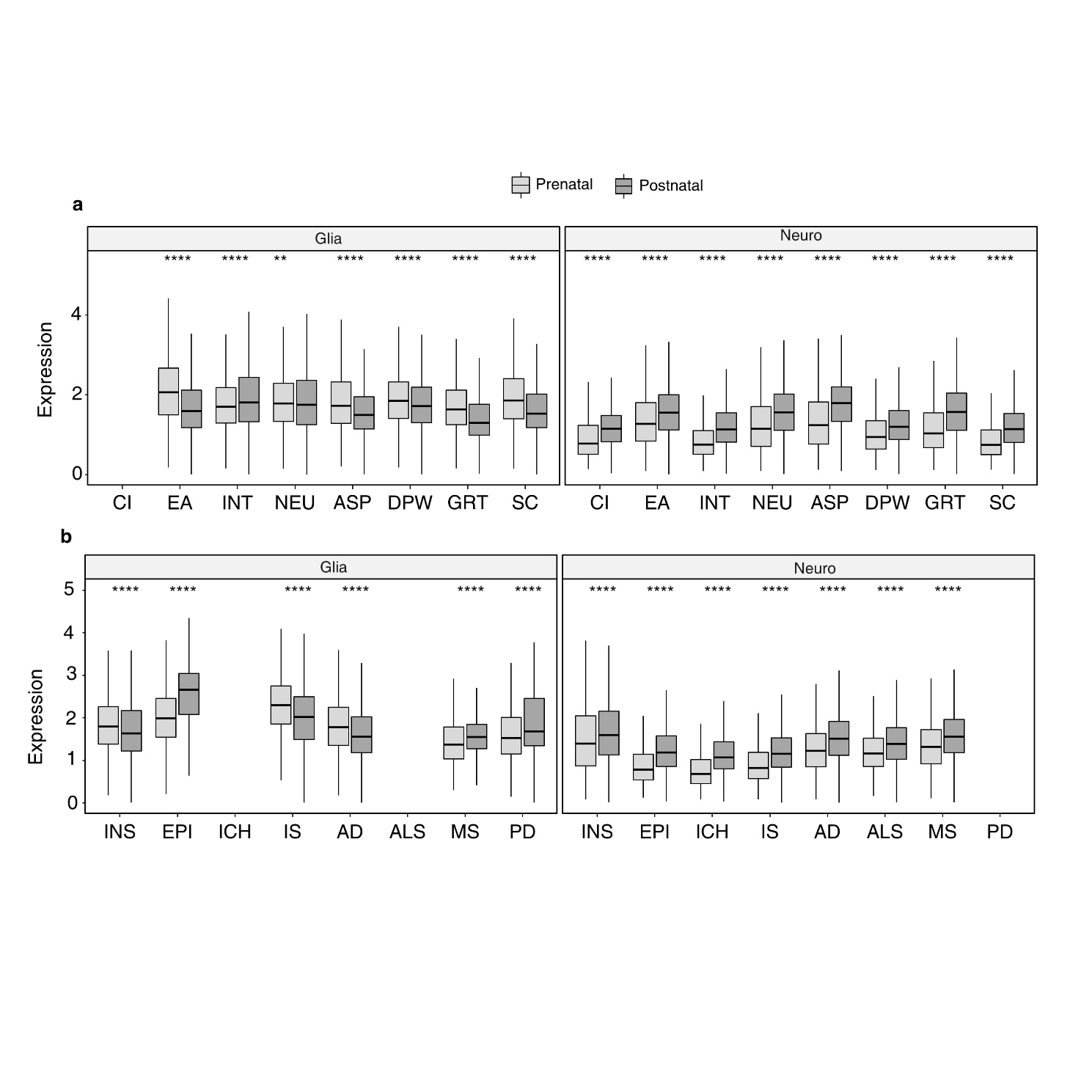

**Supplementary Fig. 9. Spatiotemporal expression levels of the causal eGenes of behavioral-cognitive phenotypes and neurological disorders**

**(a)** behavioral-cognitive phenotypes; **(b)** neurological disorders. Glia: glial cells, including astrocytes, microglia, oligodendrocytes, and oligodendrocyte precursor cells (OPCs). Neuro: neuronal cells, including inhibitory neurons and excitatory neurons.*, P < 0.05; **, P < 0.01; ****, P< 0.001.

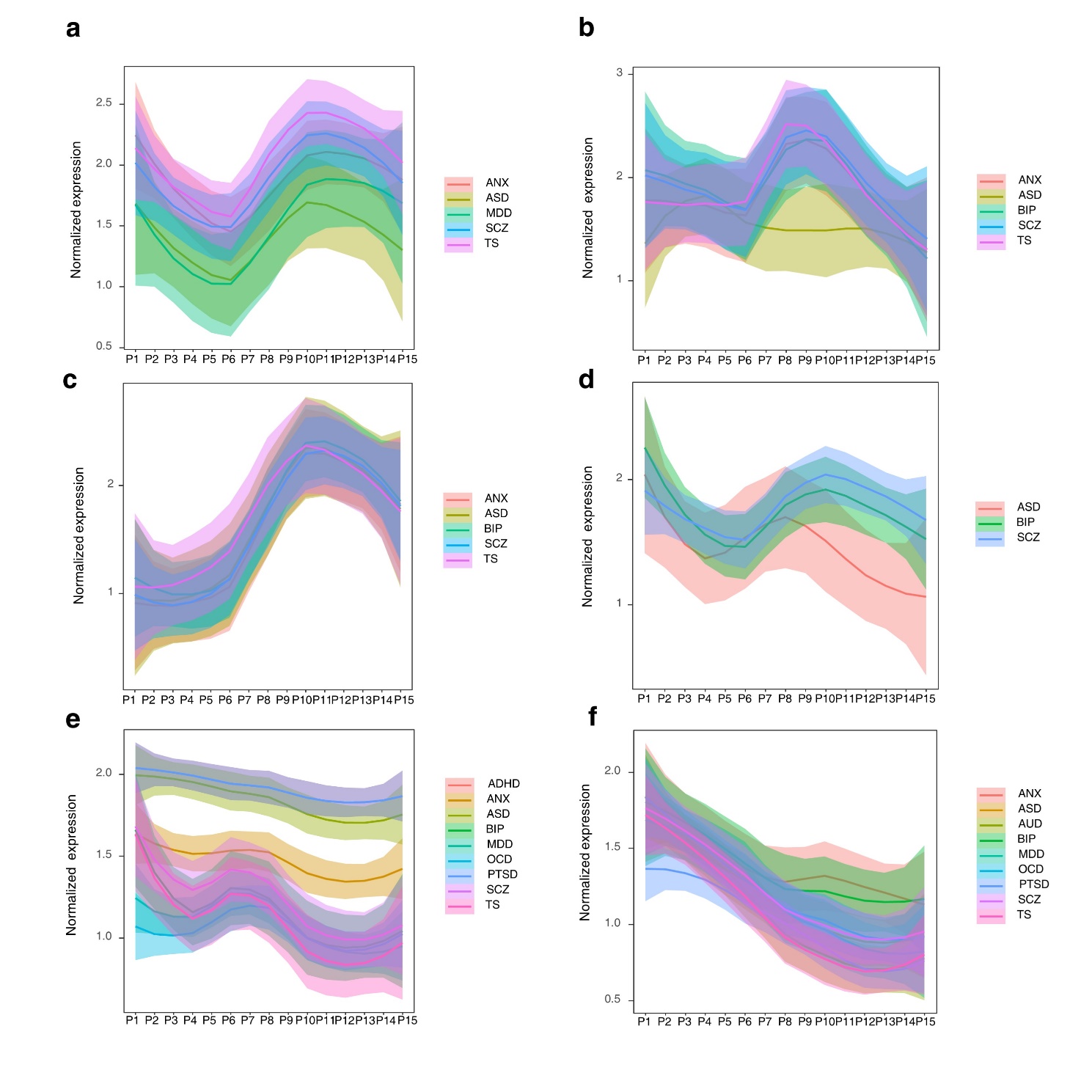

**Supplementary Fig. 10.** **Expression dynamics of the causal eGenes of psychiatric disorders from six cell types**

**(a)** astrocytes; **(b)** oligodendrocytes; **(c)** microglia**; (d)** oligodendrocyte precursor cells (OPCs); **(e)** inhibitory neurons; **(f)** excitatory neurons. For the locally estimated scatterplot smoothing (LOESS) plots, smooth curves are shown with 95% confidence intervals. P1, 4 ≤ Age < 8 PCW; P2, 8 ≤ Age < 10 PCW; P 3, 10 ≤ Age < 13 PCW; P 4, 13 ≤ Age < 16 PCW; P 5, 16 ≤ Age < 19 PCW; P 6, 19 ≤ Age < 24 PCW; P 7, 24 ≤ Age < 38 PCW; P8, 0 ≤ Age < 6 Months; P9, 6 ≤ Age < 12 Months; P10, 1 ≤ Age < 6 Years; P11, 6 ≤ Age < 12 Years; P12, 12 ≤ Age < 20 Years; P13, 20 ≤ Age < 40 Years; P14, 40 ≤ Age < 60 Years; P15, > 60 Years.

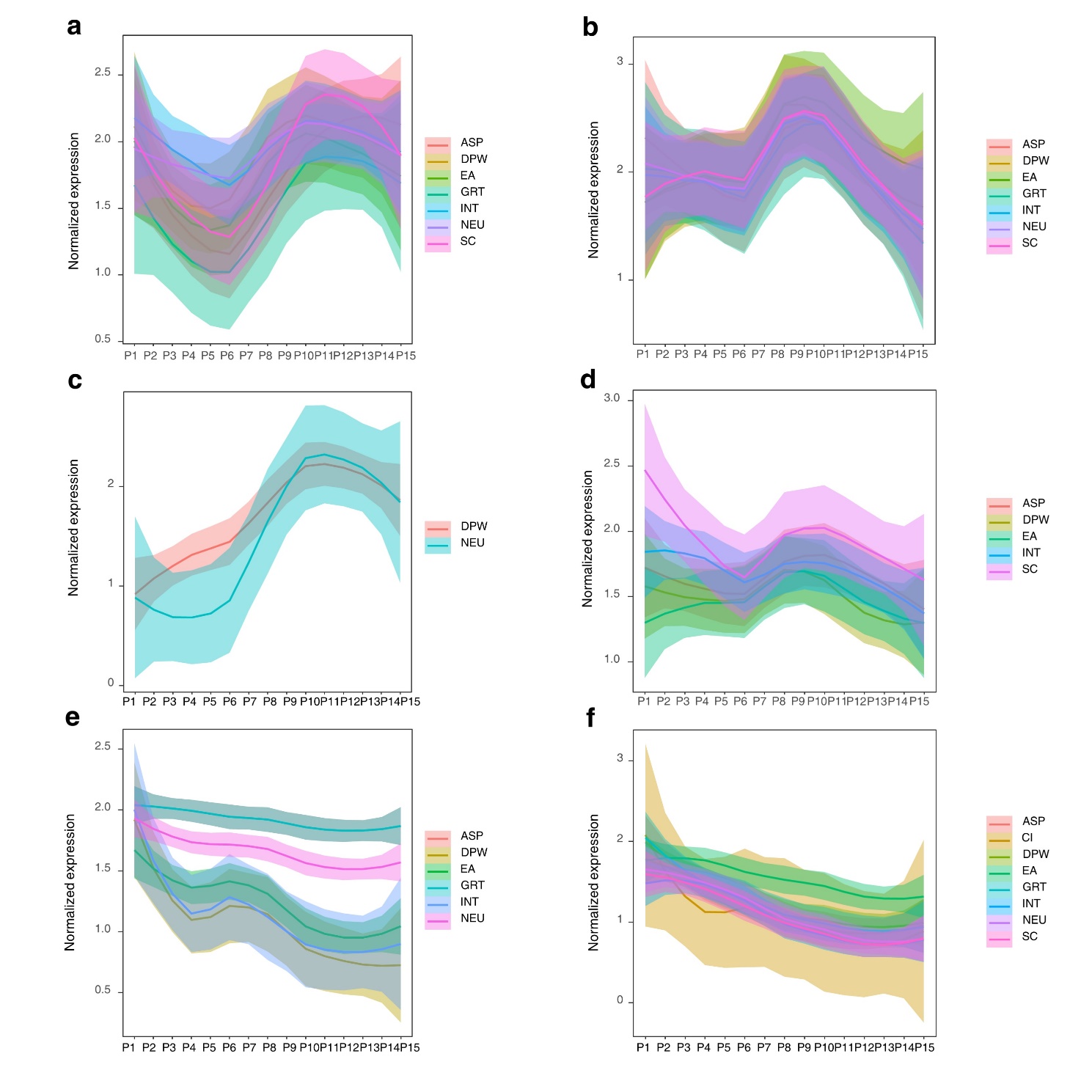

**Supplementary Fig. 11. Expression dynamics of the causal eGenes of behavioral-cognitive phenotypes from six cell types**

**(a)** astrocytes; **(b)** oligodendrocytes; **(c)** microglia; **(d)** oligodendrocyte precursor cells (OPCs); **(e)** inhibitory neurons; **(f)** excitatory neurons. For the locally estimated scatterplot smoothing (LOESS) plots, smooth curves are shown with 95% confidence intervals. P1, 4 ≤ Age < 8 PCW; P2, 8 ≤ Age < 10 PCW; P 3, 10 ≤ Age < 13 PCW; P 4, 13 ≤ Age < 16 PCW; P 5, 16 ≤ Age < 19 PCW; P 6, 19 ≤ Age < 24 PCW; P 7, 24 ≤ Age < 38 PCW; P8, 0 ≤ Age < 6 Months; P9, 6 ≤ Age < 12 Months; P10, 1 ≤ Age < 6 Years; P11, 6 ≤ Age < 12 Years; P12, 12 ≤ Age < 20 Years; P13, 20 ≤ Age < 40 Years; P14, 40 ≤ Age < 60 Years; P15, > 60 Years

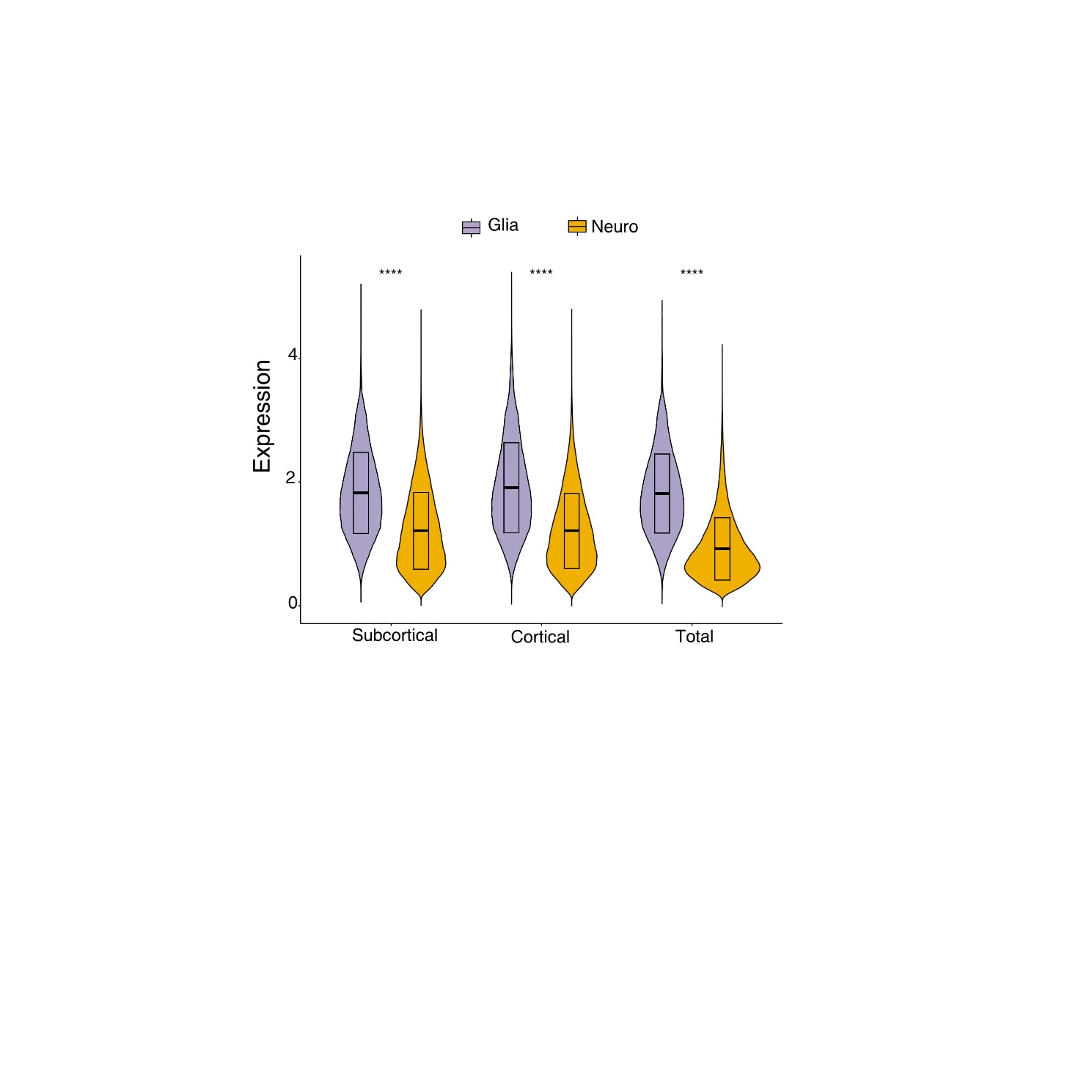

**Supplementary Fig. 12. Expression levels of the casual eGenes of 101 brain regional volumes from glial and neural cells**

The 101 brain regional volumes were categorized into three groups based on their physical positions. Glia: glial cells, including astrocytes, microglia, oligodendrocytes, and oligodendrocyte precursor cells (OPCs). Neuro: neuronal cells, including inhibitory neurons and excitatory neurons. ****, P< 0.001.

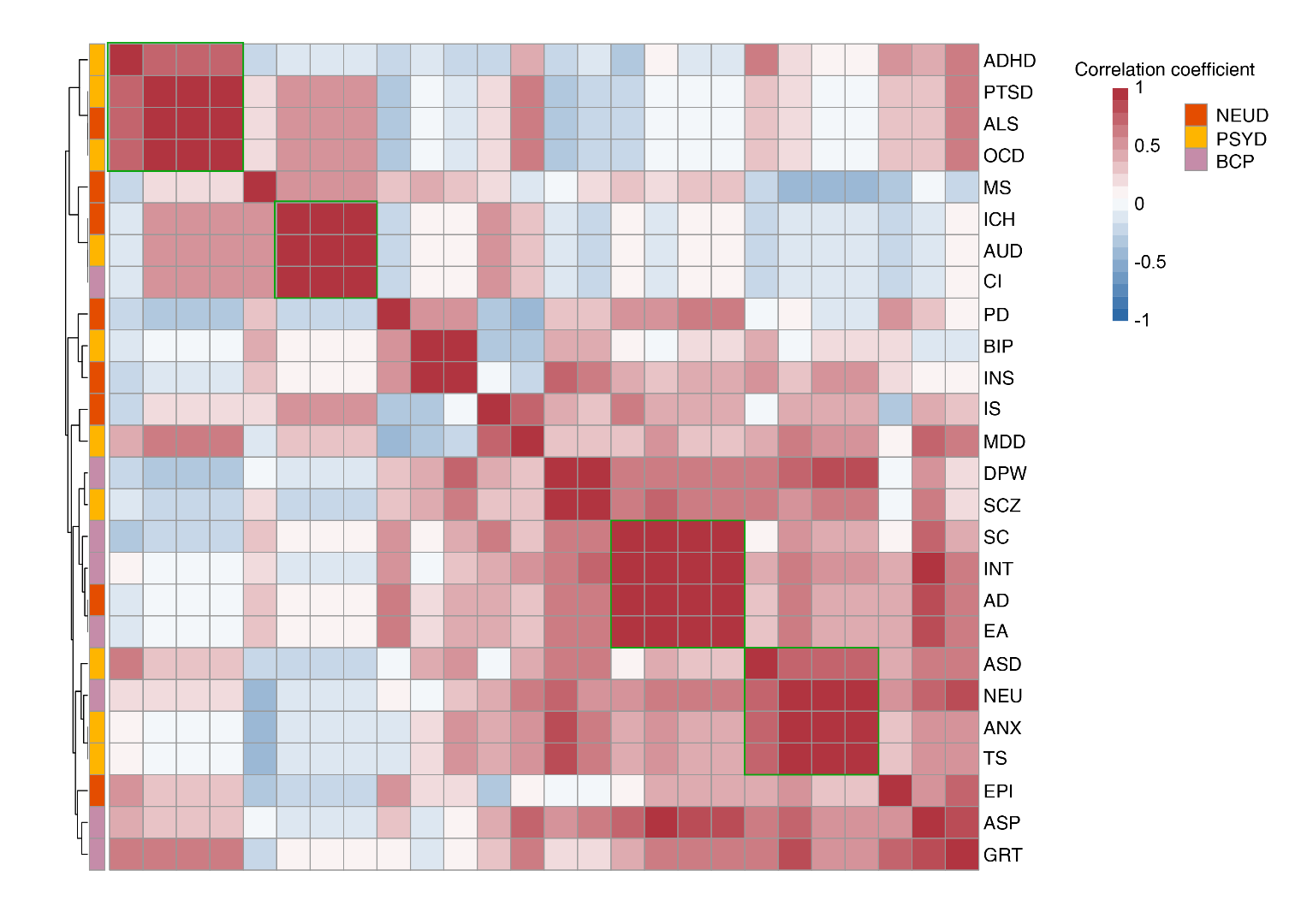

**Supplementary Fig. 13.** **Heatmap showing the associations within DBs colored by different groups**

The associations were evaluated based on the expression correlation (Spearman's correlation coefficient) of the causal eGenes from eight cell types (**Methods**), and clusters were derived using hierarchical clustering (complete linkage clustering).

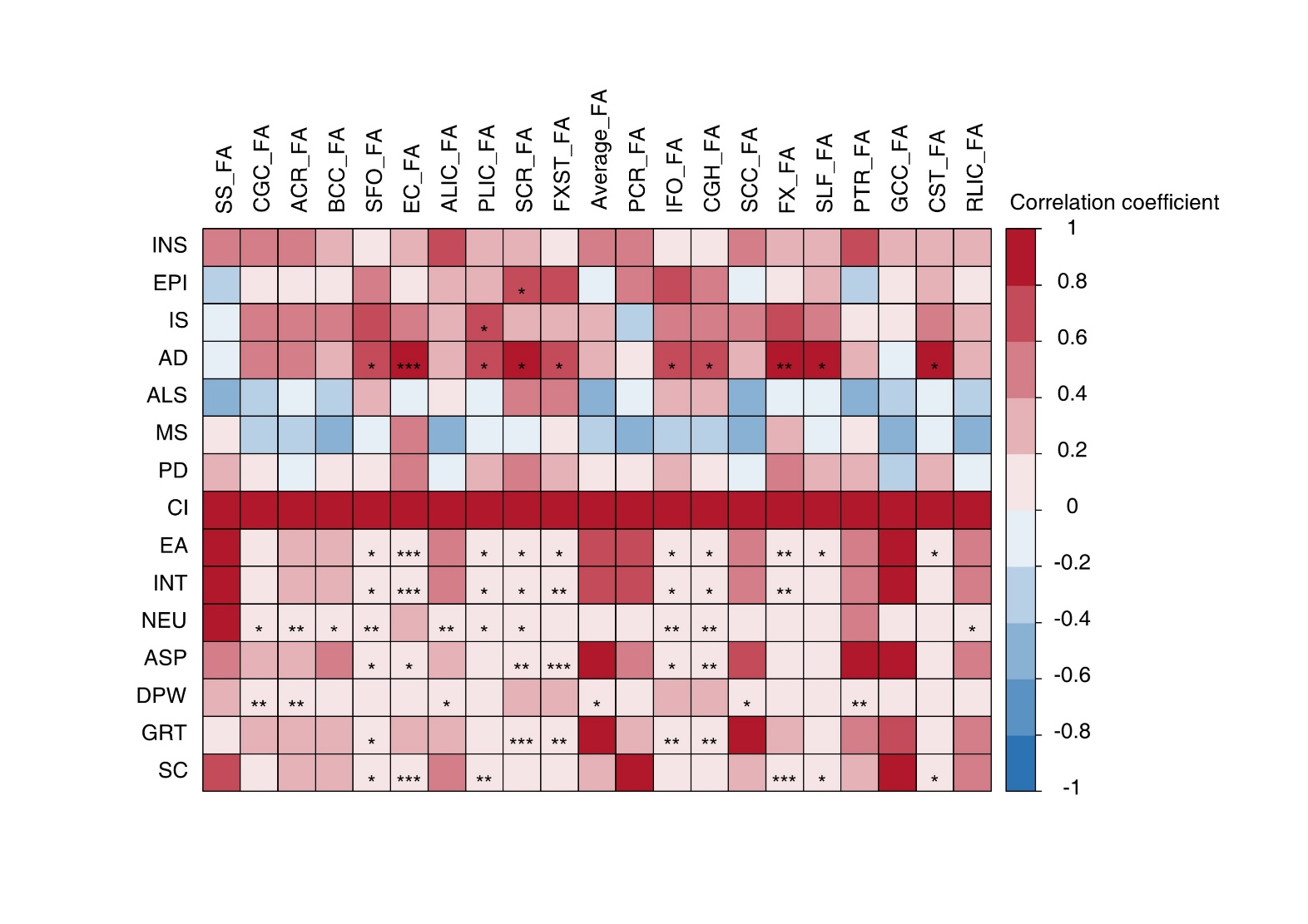

**Supplementary Fig. 14. Heatmap showing the associations between DBs and white matter microstructure phenotypes**

The associations were evaluated based on the expression correlation (Spearman's correlation coefficient) of the causal eGenes from eight cell types (**Methods**). *, FDR < 0.05; **, FDR < 0.01; ***, FDR < 0.001.

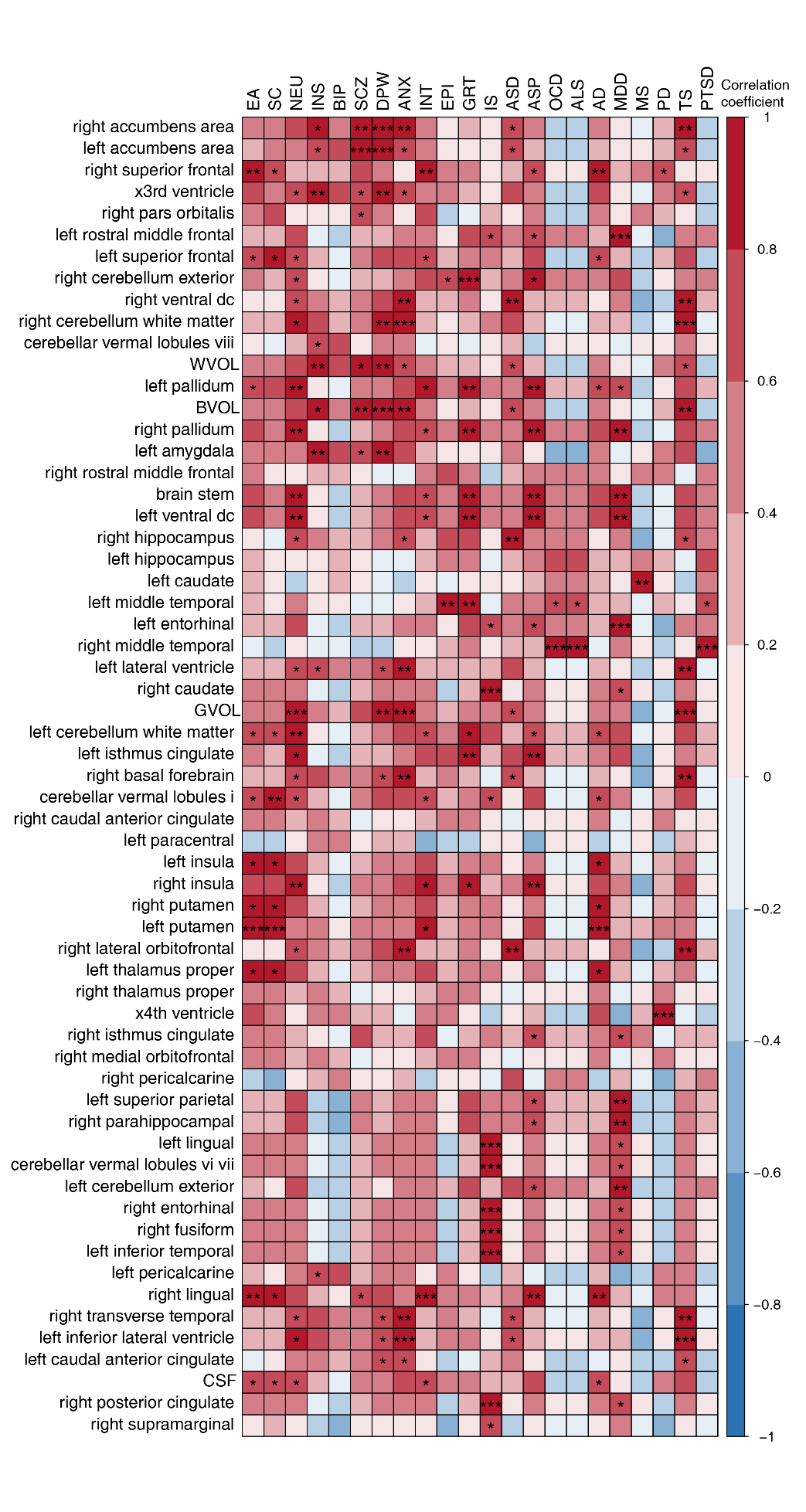

**Supplementary Fig. 15. Heatmap showing the associations between DBs and brain regional volumes**

The associations were evaluated based on the expression correlation (Spearman's correlation coefficient) of the causal eGenes from eight cell types (**Methods**). *, FDR < 0.05; **, FDR < 0.01; ***, FDR < 0.001.
